# Geospatial analysis of malaria burden in Kagera region, North-western Tanzania using health facility and community survey data

**DOI:** 10.1101/2024.06.11.24308780

**Authors:** Daniel A. Petro, Nyimvua Shaban, Sijenunu Aaron, Frank Chacky, Samwel Lazaro, Maciej F. Boni, Deus S. Ishengoma

## Abstract

Generating evidence of malaria burden in areas with confirmed artemisinin partial resistance (ART-R) is the first step before developing a response strategy to the resistance. In this study, we assessed the burden of malaria in Kagera region (with recently confirmed ART-R) by geospatial analysis using data from health facilities (HFs) and community surveys from 2017 to 2023. In 2983717/8124363 patients from HFs, rapid diagnostic test positivity rate was 36.7% (range: 0-80%) and was similar in patients aged under-fives (33.1%, range: 0-79%) and ≥5 years (33.7%, range: 3-80%). The prevalence of malaria was 10.0% (range: 0-40.5%, n=84999/ 853761) in pregnant women but it was 26.1% (3409/13065) in school children (range: 0-78.4%). We identified hotspots and coldspots, and persistently high burden in 69/192 wards, school children, and patients aged ≥5 years, providing evidence for planning and developing an ART-R response for the region.

---

Tanzania is one of the priority countries defined by the World Health Organization (WHO) as a high-burden high-impact country; it accounted for an estimated 3.2% of all malaria cases and 4.4% of deaths reported worldwide in 2022 (*1*). In Tanzania, about 96% of all cases are caused by *Plasmodium falciparum,* and the remaining are due to other species (*P. malariae* and *P. ovale*) individually or in mixed infections (*2–4*). The National Malaria Control Programme’s (NMCP) strategy is to reduce the malaria burden and progress to substantially control and eliminate using different effective interventions. The main strategies include integrated malaria vector control (mainly with insecticide-treated nets), case management, preventive therapies, and other supportive strategies (*5*). However, progress to malaria elimination is under key biological threats such as the emergence and spread of insecticide (*6*) and antimalarial drug resistance (*7*), emergence of parasites with histidine-rich protein 2/3 (*hrp2/3*) gene deletions (*8*), and invasive vector species (*Anopheles stephensi*) in some malaria-endemic countries in Africa (*1*).

Malaria transmission in Tanzania is heterogeneous with 93% of the population living in areas where transmission occurs (*5*). Regions with very low and low transmission risk are located in the central, northeastern, and southwestern parts surrounded on both sides by moderate transmission regions and high transmission in the northwestern, southern, and western parts (*9*). Various factors including the types of mosquito vectors, environmental conditions, and other human-related factors are responsible for the current pattern of malaria transmission in Tanzania (*10–12*).

Over the past four decades, malaria-endemic countries in sub-Saharan Africa have reported resistance of *P. falciparum* to all major antimalarial drugs, including chloroquine, sulfadoxine-pyrimethamine (SP) (*7*), and artemisinin derivatives and their currently used partner drugs (*13,14*). Nevertheless, artemisinin-based combination therapies (ACTs) are still highly effective against parasites of African origin (*7*). However, artemisinin partial resistance (ART-R) has been confirmed in Rwanda (*15,16*), Uganda (*17*), Tanzania (*18*) and Eritrea (*19*), and one particular genotype has only been detected in the Horn of Africa, in Ethiopia (*20*), Somalia (*21*) and Sudan (*22,23*). In addition, the mutation that originated in Rwanda has been also reported in Kenya (*24*). In Tanzania, little was known about ART-R until in 2021 when a country-wide survey through the project on molecular surveillance of malaria in Tanzania (MSMT) reported a focus in Kagera region only, with high frequency (0.077) of parasites with mutations in Kelch13 gene (*K13*) and as high as 0.22 in Karagwe District Council (DC) which are linked to ART-R in Africa (*25*). Based on WHO’s criteria (*7*), a follow-up therapeutic efficacy study conducted in 2022 confirmed the presence of ART-R in the region although the tested drugs still had high efficacy exceeding 98.0% (*18*).

As per WHO’s strategy to respond to ART-R in Africa, when resistance is confirmed, new strategies to contain, prevent, and mitigate the spread and consequences of ART-R must be considered urgently (*26*). As the first step towards the development of a response strategy, WHO recommends mapping and providing a detailed profile of the disease burden and its spread in different areas (*27*). Therefore, this study aimed to investigate the temporal trends and spatial patterns of malaria burden in Kagera region and identify areas at the highest or lowest risk of malaria transmission as an initial stage towards the development of a response to ART-R in Tanzania. The findings from this study provide the government and other stakeholders with evidence to support the designing, deployment, and implementation of interventions to monitor, prevent, and contain the spread of ART-R within the region and to other areas of Tanzania.

## Material and Methods

### Study design and site

This study utilized aggregated secondary data collected retrospectively from health facilities (HFs) and community surveys in Kagera region of north-western Tanzania. The region consists of 8 councils, 192 wards, and 734 villages covering an area of approximately 35868 square kilometers. A description of the region’s geographical boundaries and the source of its economy is provided in Appendix A.

### Data collation, management, and analysis

The data for this were downloaded from the District Health Information System – version 2 (DHIS2), a web-based software platform for reporting, analyzing, storing, and disseminating health data. This (DHIS2) is an electronic format for the paper-based Health Management Information System of the Tanzania Ministry of Health which is used by HFs to collect and report their routine data (*5*). The description of how the information is entered and maintained as well as quality control of the data is given elsewhere (*9,28*). The data used in this study included monthly malaria testing of pregnant women attending first antenatal care (ANC) clinics, symptomatic malaria patients seeking care at HFs (aged under-fives and ≥5 years) both from 2017 to 2023 and school children through school malaria parasitological surveys (SMPS) of 2015, 2017, 2019, and 2021 from the 37 selected wards in all councils of Kagera region.

Using Excel (<u>Office 2013</u>) and R software version 4.3.2: (www.r-project.org), data cleaning and preprocessing were done to ensure accuracy and consistency (Appendix B). Monthly data of total malaria tests performed and positive tests (by RDTs) reported by all HFs in the region were aggregated to provide annualized estimates. Malaria test-positivity rate (TPR) and prevalence were defined as the proportion of positive malaria tests among all malaria tests in patients aged under-fives and ≥5 years, and pregnant women, respectively.

For school children, the average prevalence for each council and ward was used. Using R software, data analysis was done for overall, per year and study group, council-wise, and at ward level, and the results presented in tables, figures or text. Python plotting tools (pandas v2.2.0, matplotlib v3.8.2, numpy v1.26.3) (Python.org) were used to show temporal trends of TPR and prevalence in the councils.

Kruskal-Wallis test was used to assess for any significant differences in malaria burden among councils, wards, and across years. Data on administrative boundaries of Tanzania at lower levels and its neighboring countries were downloaded from various sources including the Tanzanian <u>National Bureau of Statistics</u> (NBS), gadm.org, and naturalearthdata.com accessed via the R package *rnaturalearth* v1.0.1. These were used to create the study site and used for choropleth mapping, spatial autocorrelation (SA), and hotspot analysis. The spatial distribution of malaria burden in the region was mapped using R packages *ggplot2* v3.5.1 and *sf* v1.0.15.

To investigate the spatial and temporal patterns of malaria burden in the region, the Global Moran’s Index was calculated for each group and year. The value of this index ranges from −1 (negative SA) to +1 (positive SA) (*29,30*). For hotspot analysis, local spatial statistical methods were applied through a *spdep* package v1.3.3 found in R software. The description of wards and HFs included in the analysis is found in Appendix B and Figure 1, while the cut-offs applied to characterize malaria prevalence were; <1% (very low), 1 - <5% (low), 5 -<30% (moderate), and ≥30% (high) as previously described (*31,32*). For malaria TPR, the cut-offs were <5% (very low), 5-<15% (low), 15-<30% (moderate), and ≥30% (high) (Appendix C and Table 1). The formulae and illustration of how SA and hotspot analysis were conducted can be found elsewhere (*29,30*) and in Appendix D.

**Figure 1.**
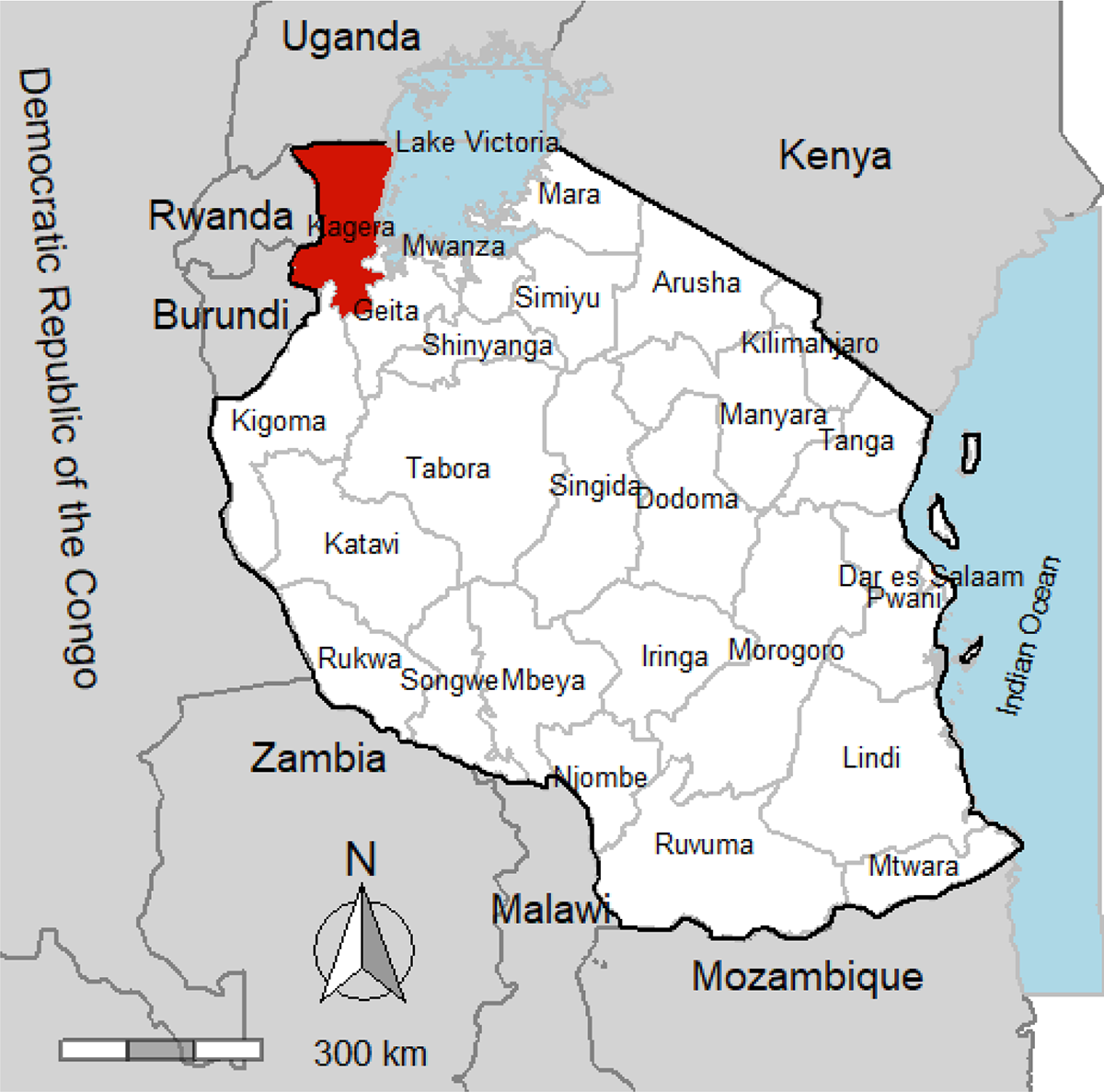
A map showing the 26 regions of Mainland Tanzania (white) and Kagera region (red), Lake Victoria and Indian Ocean (lightblue), and neighboring countries (gray).

**Table 1.** Results of global spatial autocorrelation of malaria test positivity rates in patients aged under-fives and ≥ 5 years in Kagera region.

| Group | Year | Global moran's index | z score | p value | Pattern |
| --- | --- | --- | --- | --- | --- |
| Under-fives | 2017 | 0.7281 | 12.621 | **<0.0001** | Clustered |
|  | 2018 | 0.7522 | 13.263 | **<0.0001** | Clustered |
|  | 2019 | 0.7815 | 13.771 | **<0.0001** | Clustered |
|  | 2020 | 0.7391 | 13.528 | **<0.0001** | Clustered |
|  | 2021 | 0.6745 | 12.331 | **<0.0001** | Clustered |
|  | 2022 | 0.6724 | 12.084 | **<0.0001** | Clustered |
|  | 2023 | 0.5229 | 9.440 | **<0.0001** | Clustered |
| $\geq 5$ years | 2017 | 0.6561 | 11.392 | **<0.0001** | Clustered |
|  | 2018 | 0.6852 | 12.091 | **<0.0001** | Clustered |
|  | 2019 | 0.6806 | 11.995 | **<0.0001** | Clustered |
|  | 2020 | 0.6104 | 11.197 | **<0.0001** | Clustered |
|  | 2021 | 0.6324 | 11.572 | **<0.0001** | Clustered |
|  | 2022 | 0.5779 | 10.413 | **<0.0001** | Clustered |
|  | 2023 | 0.4784 | 8.709 | **<0.0001** | Clustered |

## Results

### Reported malaria cases from Health Facilities (HFs)

In the HFs in Kagera region, 36.7% of patients (range: 0-80%; n = 2983717/8124363) with malaria-like symptoms seeking care from 2017 to 2023, had RDT-positive results. The TPRs were similar in patients aged under-fives (33.1%, range: 0-79%) and ≥5 years (33.7%, range: 3-80%), with clear geographic and temporal variations (p<0.001). Among under-fives, most of the wards in the high stratum (TPR ≥30%) were from rural areas while wards in the very low stratum (TPR <5%) came from urban areas (Appendix Table 2). Furthermore, five councils had >10% of the wards with consistently high TPR (≥30%) throughout the years. These included 6/17 (35.3%) wards in Biharamulo DC, 12/43 (27.9%) in Muleba DC, 5/22 (22.7%) in Ngara DC, 4/23 (17.4%) in Karagwe DC and 4/29 (13.8%) in Bukoba DC. In Kyerwa DC, only 2/24 (8.3%) wards had consistently higher TPRs while only one ward (Kashai in Bukoba MC) recorded the lowest TPR (<5%) (Appendix Table 4). Among patients aged ≥5 years, the pattern was similar to that of under-fives, with a large number of wards observed in the high stratum throughout the years (Appendix Table 3).

**Table 2.** Year-wise results of global spatial autocorrelation of malaria prevalence in Kagera region.

| Group | Year | Global Moran's<br>Index | z-score | p-value | Pattern |
| --- | --- | --- | --- | --- | --- |
| ANC* | 2017 | 0.3556 | 6.2041 | **<0.0001** | Clustered |
|  | 2018 | 0.5267 | 9.2397 | **<0.0001** | Clustered |
|  | 2019 | 0.5518 | 9.663 | **<0.0001** | Clustered |
|  | 2020 | 0.5383 | 9.990 | **<0.0001** | Clustered |
|  | 2021 | 0.5706 | 10.294 | **<0.0001** | Clustered |
|  | 2022 | 0.4961 | 9.275 | **<0.0001** | Clustered |
|  | 2023 | 0.3487 | 6.589 | **<0.0001** | Clustered |
| SMPS<sup>†</sup> | 2015 | 0.1688 | 0.7905 | 0.4292 | Clustered |
|  | 2017 | 0.0465 | 0.4611 | 0.6448 | Clustered |
|  | 2019 | -0.0445 | 0.1648 | 0.8691 | Dispersed |
|  | 2021 | -0.0313 | 0.1626 | 0.8709 | Dispersed |

\*ANC, Antenatal Care,
<sup>†</sup>SMPS, School malaria parasitological survey

**Table 3.** Number of wards per council classified as malaria hotspots in patients aged under-fives and ≥5 years, and pregnant women in Kagera region at 95 and 99% confidence levels.

| Group | Council | Hotspot wards | Hotspot wards |
| --- | --- | --- | --- |
|  |  | at 95% CL<sup>†</sup> (n/N, %) | at 99% CL (n/N, %) |
| Under-fives | Ngara DC* | (19/22 ,86.4) | (17/22, 77.3) |
|  | Biharamulo DC | (11/17, 64.7) | (8/17, 47.1) |
|  | Muleba DC | (15/43 ,34.9) | (12/43, 27.9) |
|  | Bukoba DC | (9/29, 31.0) | (6/29, 20.7) |
|  | Karagwe DC | (2/23, 8.7) | (1/23, 4.3) |
|  | Kyerwa DC | (1/24, 4.2) | - |
| ≥5 years | Ngara DC | (20/22, 90.9) | (16/22, 72.7) |
|  | Biharamulo DC | (9/17, 52.9) | (7/17, (41.2) |
|  | Muleba DC | (16/43, 37.2) | (13/43, 30.2) |
|  | Bukoba DC | (8/29, 27.6) | (5/29, 17.2) |
|  | Kyerwa DC | (3/24, 12.5) | (1/24, 4.2) |
|  | Karagwe DC | (1/23, 4.3) | (1/23, 4.3) |
| Pregnant women | Biharamulo DC | (13/17, 76.5) | (11/17, 64.7) |
|  | Ngara DC | (7/22 ,31.8) | (4/22, 18.2) |
|  | Muleba DC | (13/43 ,30.2) | (10/43, 23.3) |
|  | Bukoba DC | (7/29, 24.1) | (7/29, 24.1) |
|  | Karagwe DC | (4/23, 17.4) | (3/23, 13.0) |
|  | Kyerwa DC | (3/24 ,12.5) | (1/24, 4.2) |
|  | Missenyi DC | (2/20, 10) | - |

\*DC, district council, <sup>†</sup>CL, confidence level. N, number of wards within a council, n, number of wards identified as hotspots

**Table 4.**
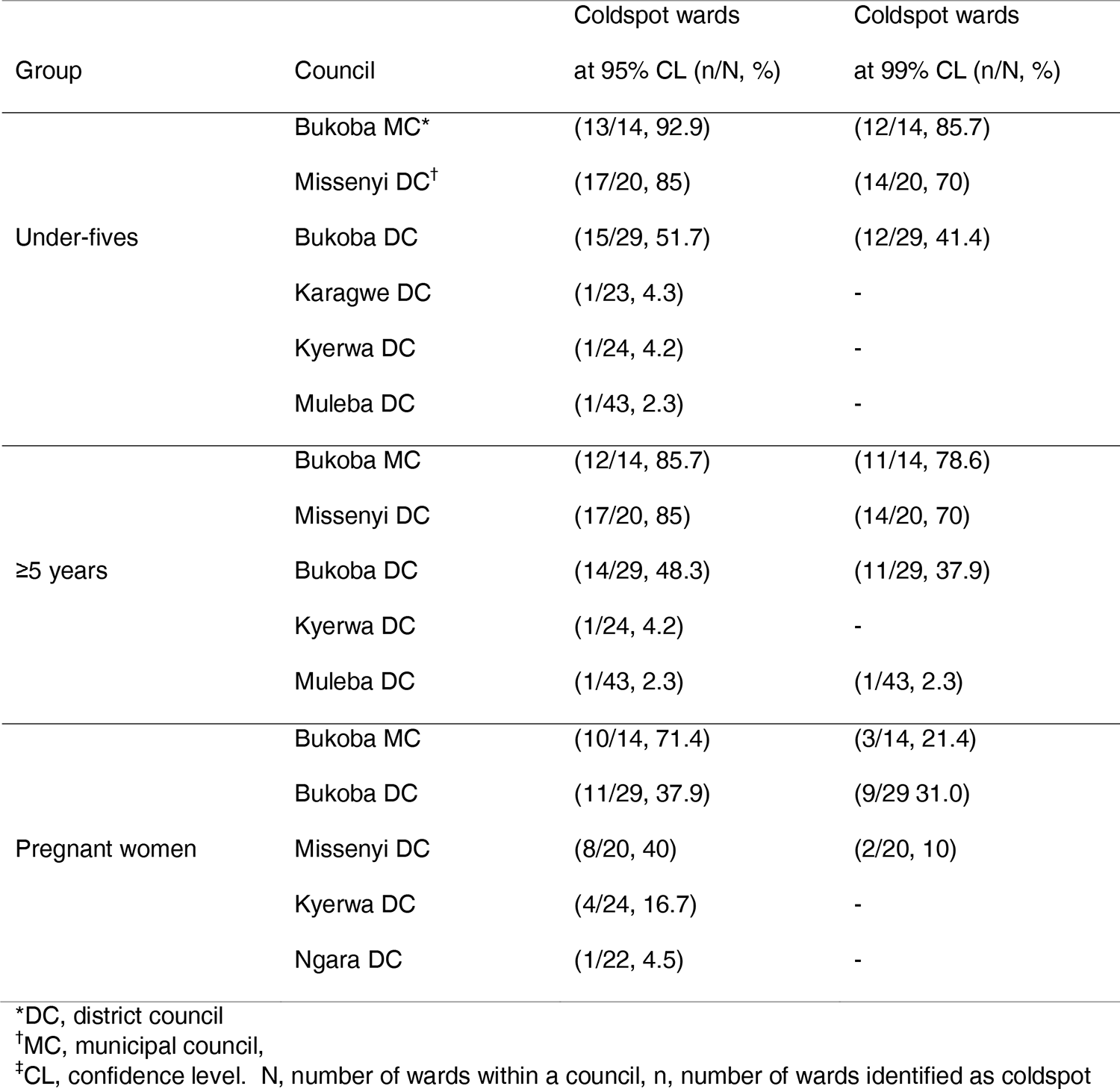
Number of wards per council classified as malaria coldspots in patients aged under-fives and ≥5 years, and pregnant women.

Similar to under-fives, the same five councils had >10% of the wards with consistently high TPRs (≥30%) over the years and these included, 7/17 (41.2%) wards in Biharamulo DC, 7/ 3 (30.4%) in Karagwe DC, 11/43 (25.6%) in Muleba DC, 5/22 (22.7%) in Ngara DC, and 4/29 (13.8%) wards in Bukoba DC. Only 1/24 (4.2%) ward in Kyerwa DC had consistently higher TPR (≥30%) (Appendix Table 4) (Figure 2B).

**Figure 2.**
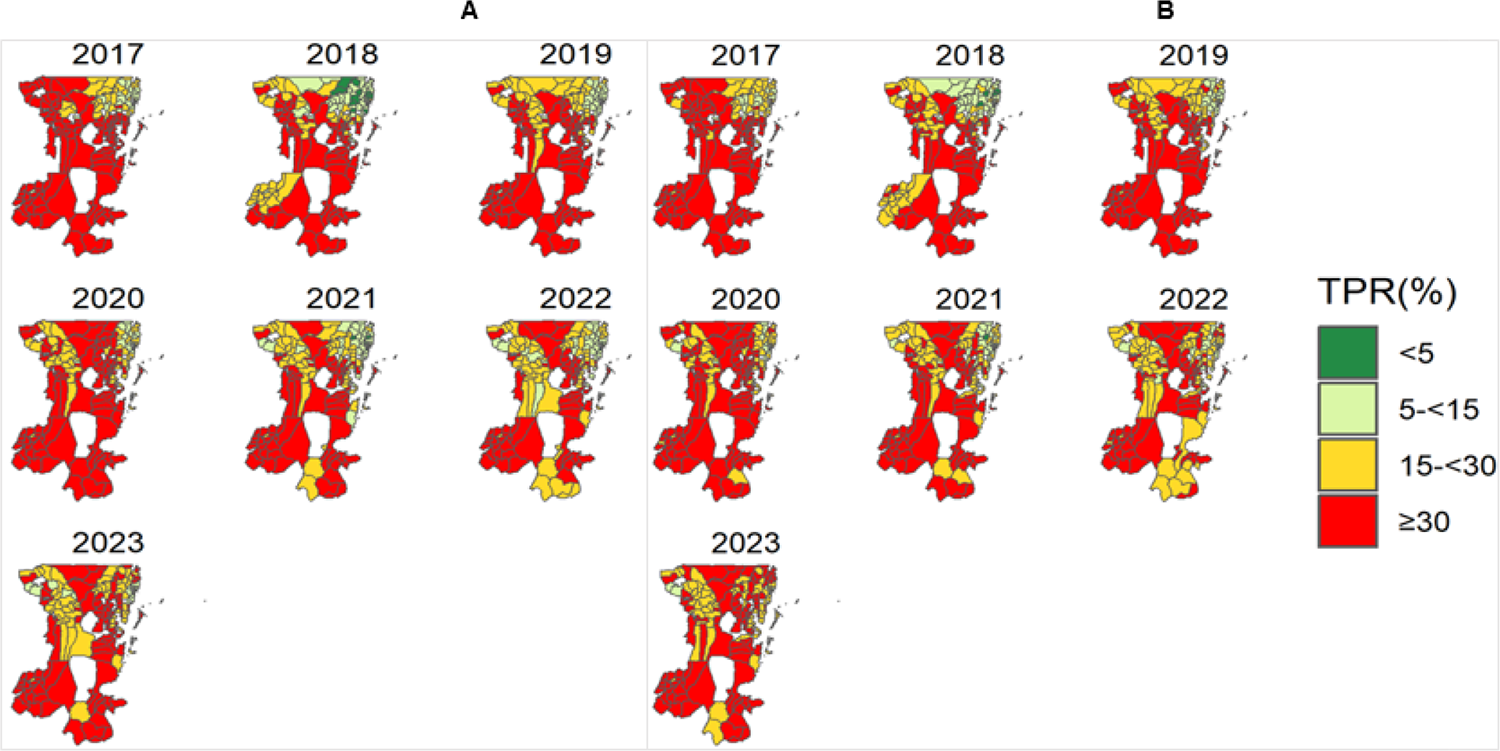
Spatial distribution of malaria test positivity rates (TPRs) in (A) under-fives (B) patients aged ≥5 years at ward level in Kagera region from 2017 to 2023. White areas are wards excluded from the analysis

### Monthly variations of test positivity rates (TPRs) at health facility level

Monthly malaria TPR was higher (≥30%) in patients aged ≥5 years compared to under-fives in all councils (in Muleba DC, Ngara DC, Biharamulo DC, Bukoba DC, and Karagwe DC) and the pattern was similar in all years, except in Bukoba MC which recorded the lowest TPR (<5%) in both age groups. In all councils except Bukoba MC and Missenyi DC, TPRs less than 20% were rarely observed (Figure 3).

**Figure 3.**
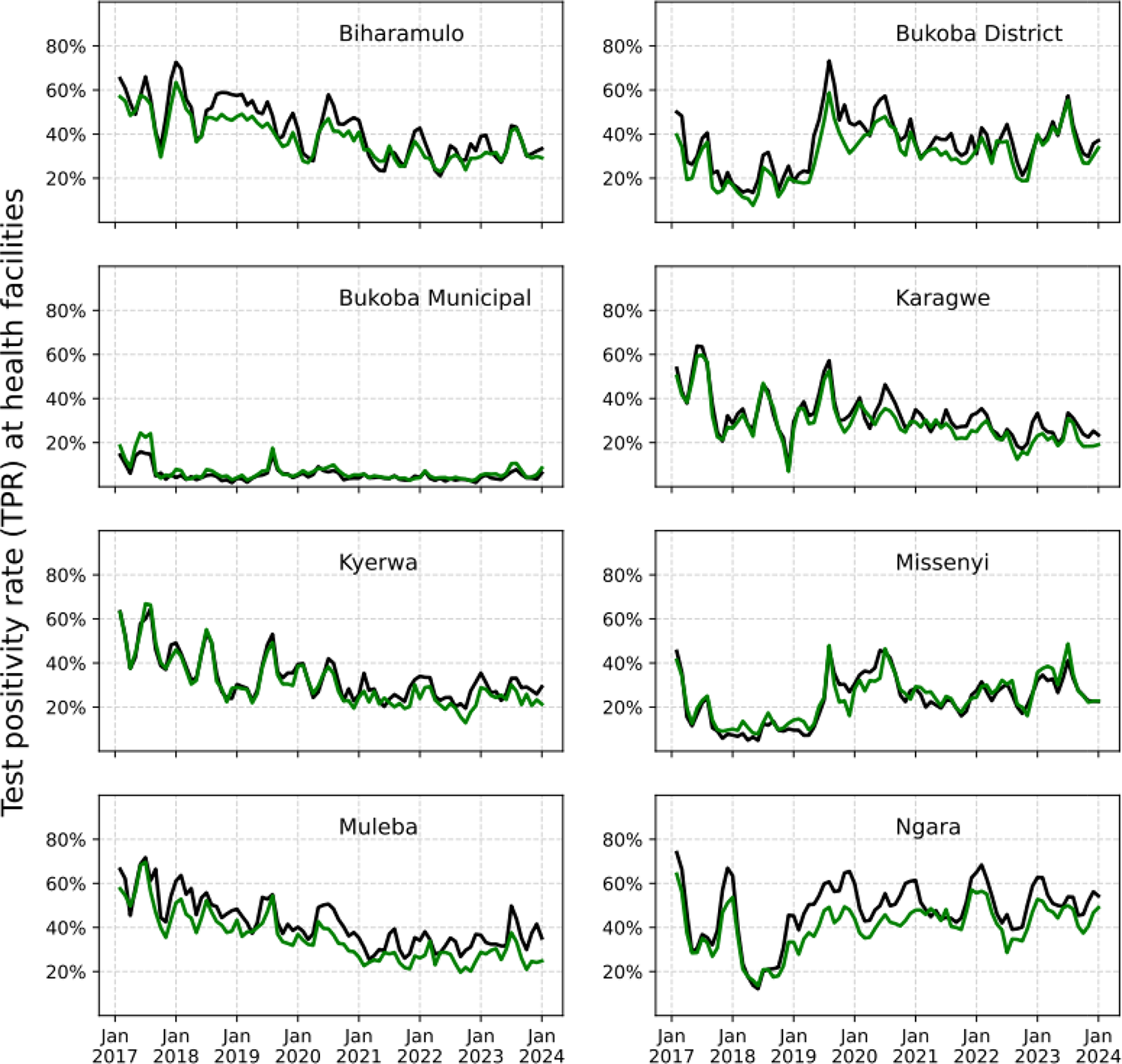
Monthly test positivity rates (TPRs) at health facility level in eight councils of Kagera region in patients aged under-fives (green) and ≥5 years (black) from 2017 to 2023.

### Malaria prevalence among pregnant women

From 2017 to 2023, the prevalence of malaria in pregnant women was 10.0% (range: 0-40.5%, n=84999/ 853761), with significant differences among wards and across years (p<0.001). The prevalence was consistently higher in Biharamulo DC, Karagwe DC, Muleba DC, and Kyerwa DC, and low in Bukoba MC (<1%) throughout the years (Figure 4).

**Figure 4.**
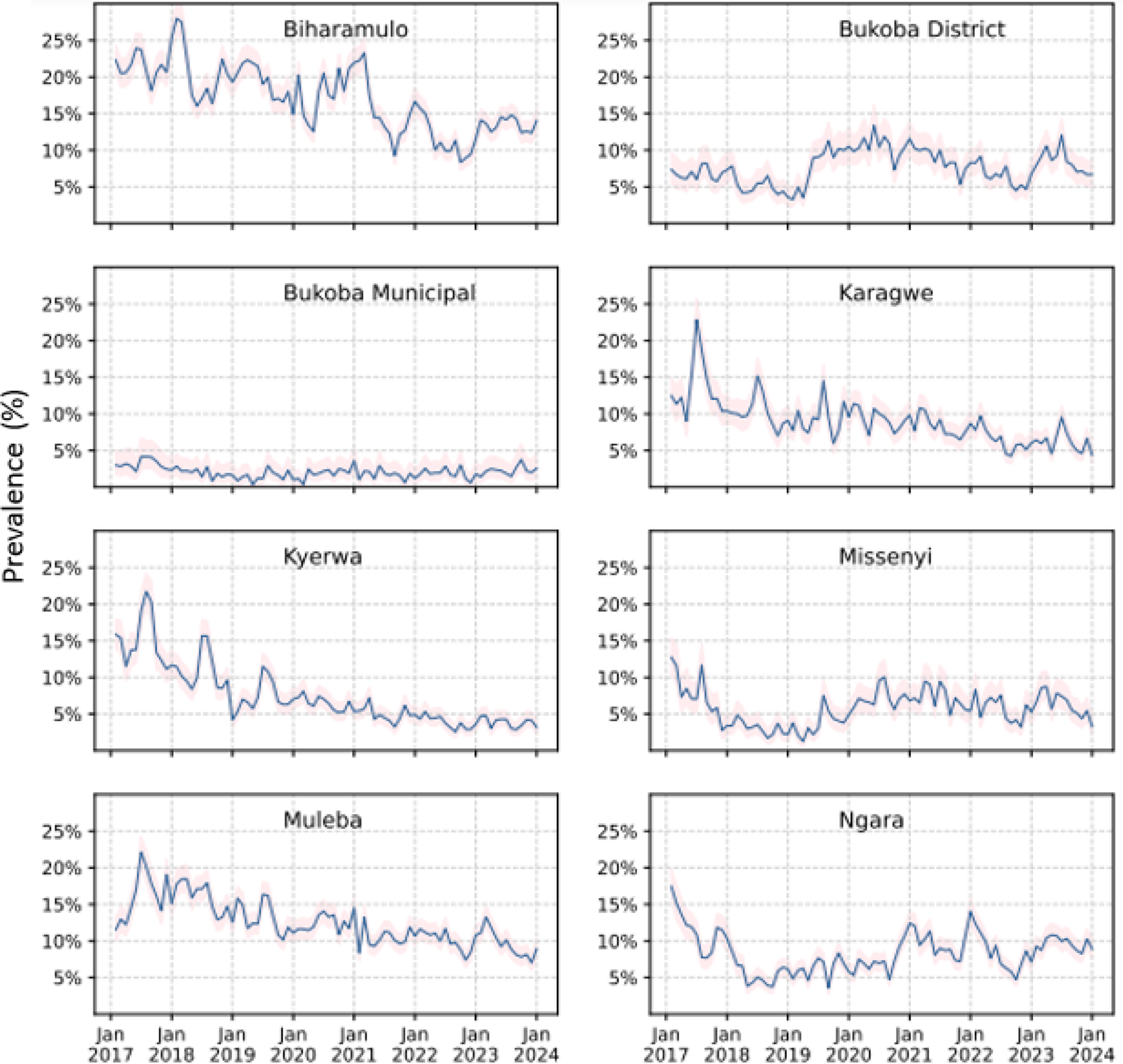
Trends of malaria prevalence in pregnant women by council, in Kagera region from 2017 to 2023. The pink-shaded regions show the Clopper-Pearson 95% confidence bands.

The largest number of wards (76.8%, n=116/151) were located in the moderate stratum while the lowest (0.6%, n=1/159) was in the high-risk stratum of prevalence (Appendix Table 5). In addition, only 3/8 (37.5%) councils had ≥5% of the wards with higher prevalence (≥30%) (including 3/17 (17.6%) wards in Biharamulo DC, 4/43 (9.3%) in Muleba DC and 2/23 (8.7%) wards in Karagwe DC) while Kyerwa DC (n=1/24, 4.2%) and Bukoba DC (n=1/29, 3.4%) had <5% of the wards with high prevalence. At least 5% of the wards from Bukoba DC (n=2/29, 6.9%) and Missenyi DC (n = 1/20, 5%) consistently recorded the lowest prevalence (Appendix Table 6) (Figure 5).

**Figure 5.**
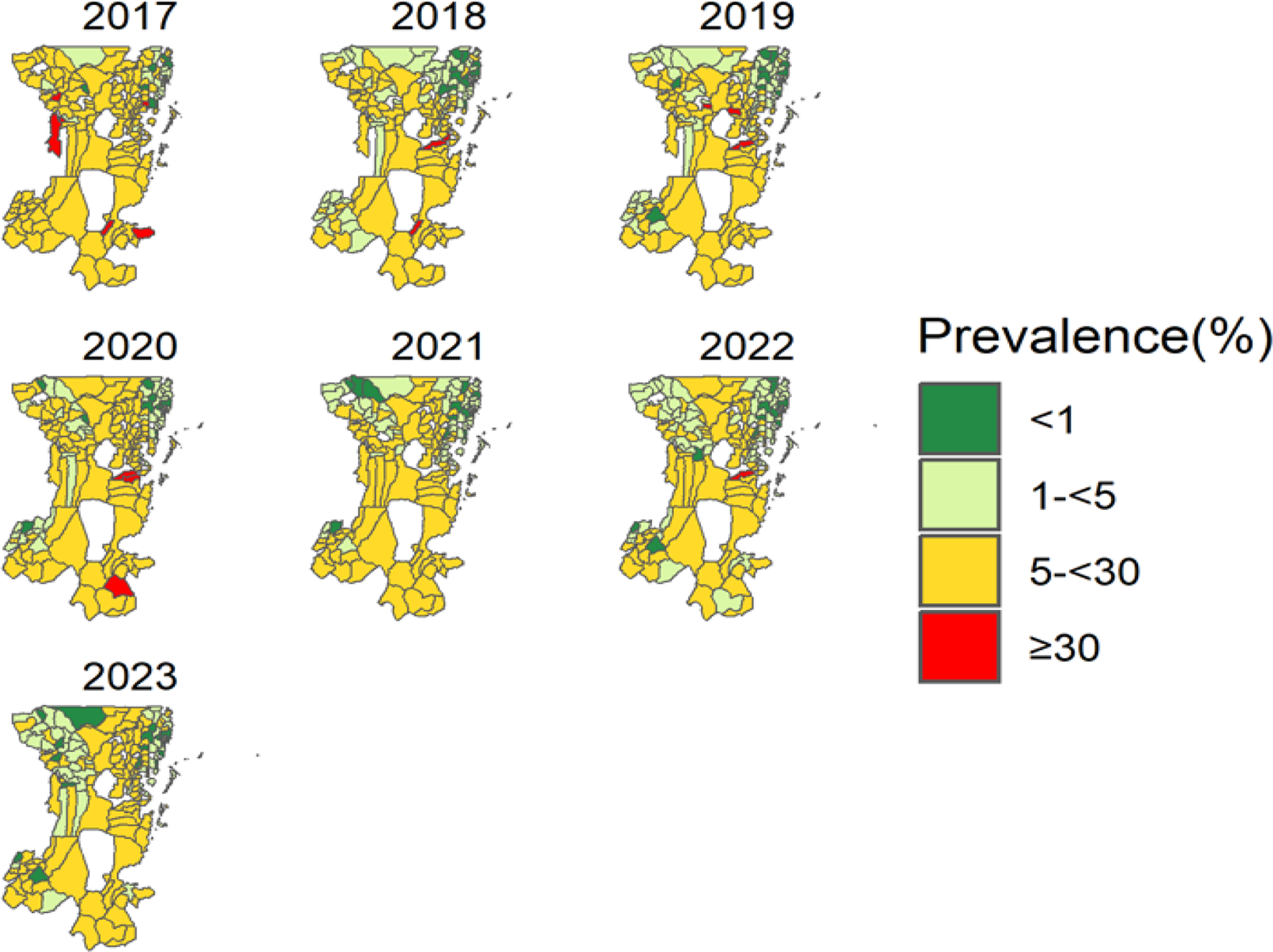
Spatial distribution of malaria prevalence in pregnant women at ward level in Kagera region from 2017 to 2023. White areas are wards excluded from the analysis.

### Malaria prevalence in school children

The prevalence of malaria in school children was 26.1% (range: 0-78.4%; n=3409/1306) (Appendix E). A large number of wards (19/34, 55.9%) were located in the high stratum (≥30%) and few wards (1/34, 2.9%) came from areas with very low stratum (<1%) (Appendix Table 7). Furthermore, two wards from Ngara DC and two from Bukoba DC had persistently higher prevalence (≥30%) throughout the study period, while no ward recorded the lowest prevalence (<1%) for all years (Appendix Table 8) (Figure 6).

**Figure 6.**
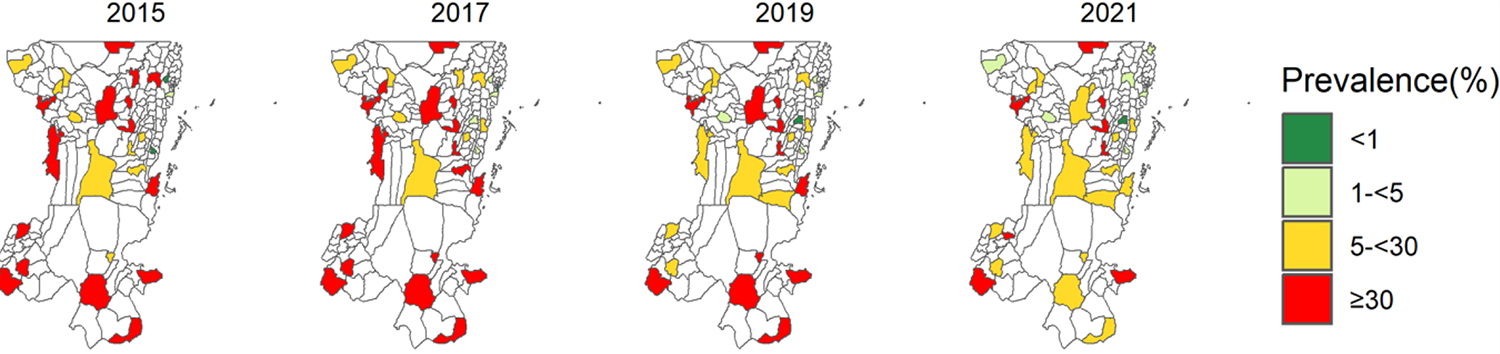
Spatial distribution of malaria prevalence in school children at ward level in Kagera region for 2015, 2017, 2019, and 2021 surveys. White areas are wards not selected for the survey or excluded from the analysis.

### Overall malaria test positivity rates at ward level

Among patients aged between under-fives and ≥5 years, a declining trend was observed in under-fives and started to shift to older patients at the same time (Appendix Table 9). In both age groups, an increasing trend was observed in 2023 (Appendix Figure 2).

### Overall malaria prevalence at ward level

Malaria prevalence was higher in school children than in pregnant women, a trend that was similar across years (Appendix Figure 3). Furthermore, prevalence in pregnant women showed a decreasing trend with minor variability before 2022 but increased by approximately 15% in 2023 (Appendix Table 9).

### Global spatial autocorrelation analysis of malaria burden

Malaria burden in the region was clustered in under-fives, patients aged ≥5 years, and pregnant women. Councils/wards with high/low burden were surrounded by other areas with high/low values as well for all years and the observed patterns were statistically significant (p<0.05) in all groups except school children (**Tables 1** and **2**).

### Hotspot analysis of malaria burden

Malaria hotspots were concentrated in the southern, central, and northwestern parts of Kagera region. Four councils (Ngara DC, Biharamulo DC, Muleba DC, and Bukoba DC) had >17% of the wards classified as malaria hotspots in patients aged under-fives and ≥5 years, and pregnant women while Karagwe DC and Kyerwa DC had <17% of wards as malaria hotspots in the same groups. The number of wards classified as hotspots of malaria in each group at 95% and 99% confidence levels (CLs) is presented in **Table 3** (Appendix Table 10).

On the other hand, malaria coldspots were situated in the northeast and few in the remaining parts of the region. Three councils (Bukoba MC, Missenyi DC, and Bukoba DC) had >20% of the wards classified as malaria coldspots in patients aged under-fives, ≥5 years, and pregnant women while Kyerwa DC and Muleba DC had <20% of the wards. The number of wards classified as malaria coldspots in each group at 95% and 99% CLs is presented in **Table 4** (Appendix Table 11).

The locations of malaria hotspots and coldspots at 95% and 99% CLs in under-fives, ≥5 years, and pregnant women respectively are indicated in Appendix Figures S4 to S6.

## Discussion

This study was conducted as an initial stage to respond to ART-R as recommended by WHO (*26*) and aimed to undertake an assessment to generate, and provide evidence of the current burden of malaria in Kagera region where ART-R was recently confirmed (*18*). We used spatial statistical and disease mapping techniques to assess the local burden of malaria and identify potential malaria hotspots in Kagera region to support NMCP’s efforts of developing a response strategy against ART-R. Using aggregated secondary data from multiple sources, we showed that over 35% (69/192) of all wards in Kagera region, mostly from rural areas, had a high malaria burden, with these wards being clustered across years. The TPR was higher in patients aged ≥5 years than under-fives and this is comparable to previous studies conducted in Tanzania (*11,12*). The findings reported here provide important evidence to support the designing and implementation of mitigation and response strategies to recently confirmed ART-R in Kagera region and Tanzania in general.

Consistent with previous studies (*12*), this study reported high temporal and spatial heterogeneity of malaria burden at council and ward levels within the region. Most of the wards, especially from rural areas, recorded a high malaria burden for most of the years; while some wards mainly in urban and mixed/rural areas recorded the lowest burden (see Appendix Tables 4,6, and 8). This finding was consistent across years and study groups possibly due to conditions supporting malaria transmission in rural compared to urban areas as previously reported (*33–35*). The higher burden of malaria in rural areas is normally attributed to several factors including the presence of favorable conditions for malaria transmission in rural areas such as stagnant water bodies that tend to create desirable breeding sites for mosquito vectors (*35*). In addition, a large proportion of poor people tend to reside in rural areas and normally live in poorly constructed houses which support higher mosquito biting rates and transmission of malaria (*12,36,37*). To reduce the burden in these areas, various interventions such as distribution of insecticide-treated nets (ITNs), house improvement strategies, bio-larviciding in areas that are surrounded by water bodies, and behavioral change and communication strategies need to be implemented, as these have been reported to significantly reduce malaria transmission and the burden (*38,39*). On the other hand, the low burden of malaria in urban areas observed in this study is consistent with previous studies (*33,35*). This is possibly due to the high socio-economic status of the urban population and, the availability and access to malaria treatment and control measures when compared to rural areas (*35*).

A significant clustered spatial pattern of malaria burden reported in patients aged under-fives and ≥5 years, and pregnant women within the region aligns with other studies conducted elsewhere (*29,30*). The reports of areas with persistently high malaria burden in the southern, central, and north-western parts of the region as reported in this study may be attributed to the presence of similar risk factors such as environmental conditions, socio-economic status, healthcare infrastructures, and the presence of suitable mosquito breeding sites that tend to create foci of malaria transmission (*10,12,40*). Hence, future interventions in the region to reduce the local disease burden need to be designed based on these results to target the areas that have been identified to have a persistently high burden of malaria.

Malaria prevalence in Kagera region was high in school children compared to pregnant women, reaching as high as 78.4% in some wards. In previous studies, it was reported that school children spend most of their time in the evening playing outside and less often use bed nets, which tends to increase their chances of mosquito bites and hence increase the risk of malaria infections (*41–43*). To reduce malaria transmission in school children, various initiatives have been undertaken by NMCP and other stakeholders to protect this risky group including the distribution of ITNs through different channels like the school net program (*5,44*). However, the prevalence of malaria infections in this asymptomatic group is still high (*12,45*). Future studies are needed to explore more reasons and factors for this recurring observation in the region and the whole country and whether this will support the spread of the reported ART-R. On the other hand, pregnant women in Tanzania are normally given two or more doses of SP for intermittent preventive treatment as well as ITNs during their first ANC visits in HFs, which might have significantly contributed to reducing the risk of malaria infections in this group (*28*). However, the increasing trend of malaria prevalence in pregnant women observed in 2023 needs further follow-up to uncover all possible reasons for this change.

The overall malaria TPR at the ward level was high (≥30%) in both under-fives and patients aged ≥5 years with minor differences. These findings are comparable to what has been reported by previous studies in Tanzania and elsewhere (*11,12*). The year-wise comparison of TPR revealed a shift of TPR from under-fives to ≥5 years, similar to previous studies (*46*). This may be attributed to various efforts that have been undertaken by NMCP to implement effective malaria interventions in vulnerable groups like under-fives while these interventions may not have been widely considered in older age groups (*11,28*). This might have led to the shift of malaria infections to individuals aged ≥5 years, with the peak of malaria prevalence reported in children aged between 5 and 15 years (11). In addition, malaria TPR in under-fives and ≥5 years showed an increasing trend in 2023. An in-depth analysis to uncover possible reasons for these epidemiological changes in the region is required so that appropriate measures can be planned, now that ART-R has been confirmed.

Malaria transmission in Kagera region was observed throughout the year with some of the months recording high values of TPR compared to others. This indicates that malaria transmission in the region was not seasonal; an observation that can signify the presence of other factors like population movements, changes in malaria control, or unexpected increases/decreases in breeding sites or mosquito populations (*12,47,48*). Causal effects for these factors are in general difficult to establish, future studies are needed to explore and uncover the key drivers of persistent malaria transmission in this malaria-endemic region in the country.

The findings presented here had some limitations that may be addressed in future studies. Since this study included HFs with a reporting rate of 50% or more, future studies may consider including all HFs during analysis and use different geostatistical methods to account for monthly missing data, differences in utilization of HFs, treatment-seeking behaviors, and distance to HFs. In addition, the HF data did include personal information such as age and sex, and this limited additional analysis. For instance, previous studies showed a high burden of malaria in school children (aged 5 - 15 years) but this could not be teased out because all patients aged ≥5 years were put in one group. Future studies may also include populations at risk and use incidence to track changes in the disease burden within the region.

Overall, malaria infections in the region were higher in most of the wards from rural areas compared to urban areas, among school children compared to pregnant women, and among patients aged ≥5 years than in under-fives. The observed heterogeneity of malaria burden underscores prioritization of Kagera region for additional malaria control mainly due to the currently circulating *P. falciparum* parasites with *K13* 561H genotypes in the region (*18*). As early and preemptive action is preferred to late response (*49*), identifying specific resistance containment strategies (*50*) for 2024 and 2025 – in addition to general malaria control strategies– will be critical for slowing down the spread of artemisinin-resistant parasites from Kagera region to other parts of Tanzania.

## Biographical Sketch

Mr. Petro is a doctoral student at the University of Dar es Salaam, Dar es Salaam. His research interests focus on malaria epidemiology.

## Data Availability

Data used in this study are not publicly available and were obtained with a request from MoH through NMCP. Restrictions apply to the availability of this data and permission can be obtained with a reasonable request from the Permanent Secretary - Ministry of Health of Mainland Tanzania.

## Acknowledgment

The authors would like to thank the MoH through NMCP, and all members of the Department of Mathematics, University of Dar es Salaam, and MSMT project team for their participation and invaluable support to make this work complete.

This study was partially funded by the National Institute for Medical Research (NIMR) under the MSMT project. The MSMT project is funded in whole, by the Bill & Melinda Gates Foundation [grant number INV 02202]. Under the grant conditions of the Foundation, a Creative Commons Attribution 4.0 Generic License has already been assigned to the author of the accepted Manuscript version that might arise from this submission.

PAP and DSI designed the study, and SA, FC and SL supported the acquisition and cleaning of the data. DAP conducted the initial analysis under the supervision of DSI and NS. DSI, NS, and MFB were involved in the interpretation of the results. DAP and MFB performed a final analysis of the data under the guidance of DSI, and DAP drafted the initial manuscript. All co-authors contributed to the revision of the initial draft and subsequent versions of the manuscript. All authors read and approved the final manuscript, and declared no competing interest.

## Appendices

**Appendix A.** Description of the study site

**Appendix B.** Administrative units and health facilities in Kagera region that were included in the analysis

**Appendix C:** Stratification of malaria burden in Kagera region

**Appendix D:** Description of spatial autocorrelation analysis

**Appendix E:** Description of wards and schools selected for school malaria parasitological surveys

**Appendix Figure 1.** Descriptive summary of the wards and health facilities included in the analysis

**Appendix Figure 2** Comparison of malaria test positivity rates in Kagera region from 2017 to 2023 based on the data from health facilities

**Appendix Figure 3.** Comparison of malaria prevalence in Kagera region from 2017 to 2023 based on the data from pregnant women and school children

**Appendix Figure 4** Locations of hotspots and coldspots of malaria in under-fives at ward level in Kagera region from 2017 to 2023

**Appendix Figure 5** Locations of hotspots and coldspots of malaria in patients aged ≥5 years at ward level in Kagera region from 2017 to 2023

**Appendix Figure 6.** Locations of hotspots and coldspots of malaria in pregnant women at ward level in Kagera region from 2017 to 2023

**Appendix Table 1.** Cut-offs adopted to categorize councils/wards in Kagera region into different risk strata

**Appendix Table 2.** Number of wards by year in each stratum of malaria test positivity rates in under-fives in Kagera region from 2017 to 2023

**Appendix Table 3.** Number of wards by year in each stratum of malaria test positivity rates in patients aged ≥ 5 years in Kagera region from 2017 to 2023

**Appendix Table 4.** Ward names by council that had a high, moderate, low, and very low malaria test positivity rates in under-fives and ≥5 years from 2017 to 2023, Kagera region

**Appendix Table 5.** Number of wards by year in each stratum of malaria prevalence in pregnant women in Kagera region from 2017 to 2023

**Appendix Table 6.** Ward names by council that had a high, moderate, low, and very low malaria prevalence in pregnant women from 2017 to 2023, Kagera region

**Appendix Table 7.** Number of wards by year in each stratum of malaria prevalence in school children in Kagera region from 2017 to 2023

**Appendix Table 8.** Ward names by council that had a high, moderate, low, and very low malaria prevalence in school children from 2017 to 2023, Kagera region

**Appendix Table 9.** Overall malaria burden at ward level in Kagera region by years and study groups from 2015 to 2023

**Appendix Table 10.** Names of hotspots of malaria burden at ward level in Kagera region at 95% and 99% confidence levels from 2017 to 2023

**Appendix Table 11.** Names of coldspots of malaria burden at ward level in Kagera region at 95% and 99% confidence levels from 2017 to 2023

