## Appendix for "Geospatial analysis of malaria burden in Kagera region, North-western Tanzania using health facility and community survey data"

**Appendix A. Study site**

This study covered Kagera region which has eight councils; namely Bukoba Municipal Council (MC, with 14 wards), Bukoba District Council (DC, with 29 wards), Biharamulo DC (17 wards), Karagwe DC (23 wards), Kyerwa DC (24 wards), Muleba DC (43 wards), Missenyi DC (20 wards), and Ngara DC (22 wards) (*1,2*). According to the 2022 national census, the region had an estimated population of 2989299 people (1530019 females and 1459280 males), with an average household of 4.3 people, 2.0 population growth rate/annuma, and a sex ratio of 96 males per 100 females (*2*). The region is bordered by Uganda in the north, and Burundi, and Rwanda in the west, and the Tanzanian regions of Kigoma and Geita in the south. To the east, the region is bordered by Lake Victoria which separates Kagera from Mwanza and Mara regions (*1*). In addition, the Kagera River forms the region’s border with Rwanda to the west and Uganda to the north, forming the biggest valley in Africa with high human movement and substantial socio-economic activities (*3*) (see Fig. 1).

The Kagera region experiences a bi-modal rainfall pattern (between March and June and from August/September to December) with an annual average ranging from 600 to 2000 millimeters (*4,5*). The driest period is from mid-June to mid-August and the second short dry season extends from January to March with average daily temperatures of 21^0^C (range: 12^0^C to 34^0^C). About 28% of the region is covered by water that comes from lakes Victoria, Ikimba, and Burigi; and big rivers such as Ngono and Kagera as well as many small perennial rivers making the region have a large volume of free-flowing fresh water throughout the year (*1*). The economy of the region depends on agriculture with coffee and bananas as the leading cash and food crops respectively, together with other activities like tourism, fishing, hunting, trading, and mining (*1*)

**Appendix B. Administrative units and health facilities in Kagera region**

In Kagera region, there are eight councils with 192 wards which have been defined according to the 2022 national census and were considered in this analysis (Appendix Figure 1). Of these, 158 (82.3%) are rural wards while 20 (10.4%) are classified as urban/rural (mixed) areas, and only 14 (7.3%) are urban wards. The number of wards per council ranges from 14 (in Bukoba MC) to 43 (in Muleba DC), depending on the council size (*2*). As of 2022, the region had 381 health facilities (HFs) which include one regional referral hospital, 21 (5.5%) hospitals, 59 (15.5%) health centres and 301 (79%) dispensaries (*2*). The number of HFs in each of the councils were; 35 (in Biharamulo DC), 31 (Bukoba MC), 44 (Bukoba DC), 49 (Karagwe DC), 39 (Kyerwa DC), 37 (Missenyi DC), 57 (Muleba DC), and 63 (in Ngara DC). In this study, data from 355 (93.2%) HFs located in 165 (85.9%) wards were downloaded from the DHIS2 and used for the analysis. Duplicates, blank reports, and HFs with no tests performed by rapid diagnostic tests (RDTs) in any of the study periods were removed from the data. Missing data in facilities with complete reports were replaced by zeros, as DHIS2 cannot distinguish between zeros and missing data. After excluding HFs without any report and facilities with no reporting rate of 50% or more, 162 (84.4%) wards consisting of 330 (92.9%) HFs were retained and later included in the analysis. Each ward depending on its size consisted of between one to 6 HFs that served the surrounding village populations.

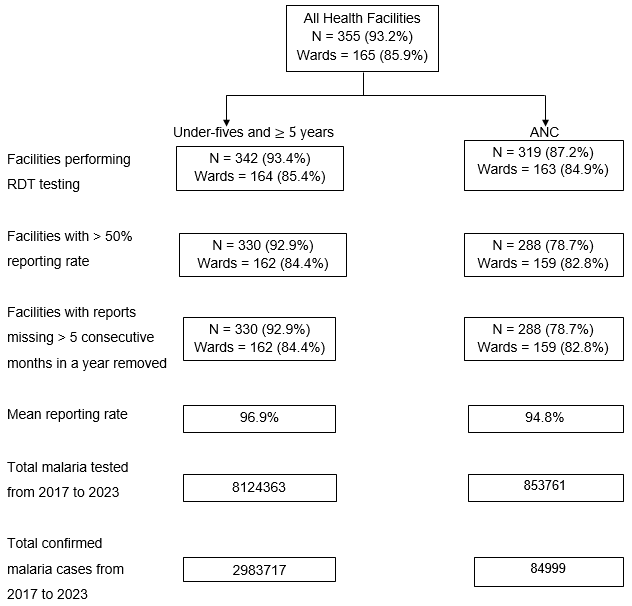

**Appendix Figure 1**. Descriptive summary of the wards and health facilities included in the analysis.

ANC = antenatal care; RDT = rapid diagnostic test.

**Appendix C. Stratification of malaria burden in Kagera region**

Although the classification of transmission settings in malaria-endemic regions remains largely arbitrary (*6*), in Tanzania, the National Malaria Control Programme (NMCP) employs various criteria to classify areas based on several factors, including parasite prevalence, transmission intensity, geographical features, and other epidemiological indicators. While exact cut-offs may vary over time based on updated data and strategies, NMCP has been using the classification of prevalence in school children (PfPR_5-16_) as a gold standard in guiding the selection of appropriate cut-offs to classify malaria risk areas in Tanzania (*7*). The classification includes both very low (PfPR_5-16_ < 1%) and high (adapted to be a PfPR_5-16_ ≥30%). Within this range, two additional groups have been considered: low (PfPR_5-16_ 1 - 5%) which provides a pre-very low classification to mitigate against the risks of misclassifying very low areas, and moderate prevalence (PfPR_5-16_ 5 - 30%) (Table 1). For malaria TPR, the cut-offs used include <5% (very low), 5-<15% (low), 15-<30% (moderate), and ≥30% (high) (*8,9*). This study used the same cut-offs recommended by NMCP to characterize malaria burden across councils and wards in the Kagera region in each of the study groups.

**Appendix Table 1.** Cut-offs (%) adopted to categorize councils/wards into different risk strata (*8,9*)

| Malaria burden | Very low | Low | Moderate | High |
| --- | --- | --- | --- | --- |
| Prevalence | <1 | 1-<5 | 5-<30 | ≥30 |
| Test positivity rate | <5 | 5-<15 | 15-<30 | ≥30 |

**Appendix D. Spatial autocorrelation analysis**

The spatial patterns of malaria burden were examined through global spatial autocorrelation (SA) analysis (Global Moran’s index) which highlights whether the patterns are clustered, dispersed, or random (see Tables 1 and 2). This statistical test compares the values of nearby places to the overall mean, and when nearby units across the entire region have comparable values both below or above the mean, a positive SA (clustered) is indicated whereas, a negative SA (dispersed) is shown by the statistic when neighboring units have diverging values (*10,11*). The analysis involved calculating Moran’s I Index value and both a *z*-score and *p*-value to evaluate the significance of that index for each year. In calculating Moran’s I index, Queen’s contiguity method was applied to construct a spatial weights matrix, a matrix that represents the spatial relationship between subunits that together comprise the whole study area. It consists of 1 if two subunits are neighbors and zero otherwise. In this case, two districts/wards were considered neighbors if they shared at least a vertex or an edge (*12*). Using a *spdep* package from R programming software *v*4.3.2, the patterns and SA results were evaluated for each year together with their corresponding *z*- and *p*-values, and the results are presented in Tables 1 and 2.

Since the results of SA indicated the presence of significant clustered patterns in under-fives and ≥5 years, and pregnant women only, local SA (hotspot analysis) was then performed to identify districts/wards with statistically significant hotspots and coldspots in the region. Here, two forms of analysis were done to investigate whether each subunit was surrounded by other units with similar (High-High or Low-Low) or dissimilar (High-Low or Low-High) values of malaria prevalence/TPR and if there were any significant associations between areas with high or low values (*10,11*). For any district/ward to be a statistically significant hotspot, it should have a high value of malaria prevalence or TPR and be surrounded by other districts/wards with high values as well. Hotspot analysis identified statistically significant hotspots and coldspots through the Local Moran’s and Getis-Ord (G­i*) statistics for each feature (district/ward) in the dataset. Based on these analyses, a *z*-score for each locality in the Kagera region was returned, where significant positive *z*-scores referred to the more intense clustering of high values (hotspot) and significant negative z-scores indicated more intense clustering of low values (coldspot) (*10*). More description of how SA and hotspot analysis are done can be found elsewhere (*11,12*). The significance level chosen in this study was *p=0.05.*

**Appendix Table 2.** Number of wards by year in each stratum of malaria test positivity rates in under-fives in Kagera region from 2017 to 2023

| Strata/Year | 2017 | 2018 | 2019 | 2020 | 2021 | 2022 | 2023 |
| --- | --- | --- | --- | --- | --- | --- | --- |
| High | 103 | 67 | 85 | 92 | 65 | 62 | 83 |
| Moderate | 28 | 39 | 41 | 44 | 53 | 59 | 54 |
| Low | 23 | 36 | 30 | 21 | 36 | 33 | 21 |
| Very low | 2 | 16 | 2 | 4 | 8 | 7 | 3 |
| Total | 156 | 158 | 158 | 161 | 162 | 161 | 161 |

**Appendix Table 3.** Number of wards by year in each stratum of malaria test positivity rates in patients aged ≥ 5 years in Kagera region from 2017 to 2023

| Strata/Year | 2017 | 2018 | 2019 | 2020 | 2021 | 2022 | 2023 |
| --- | --- | --- | --- | --- | --- | --- | --- |
| High | 103 | 65 | 89 | 98 | 76 | 70 | 88 |
| Moderate | 36 | 46 | 43 | 43 | 56 | 67 | 62 |
| Low | 17 | 41 | 24 | 19 | 26 | 20 | 11 |
| Very low | - | 6 | 2 | 1 | 4 | 4 | - |
| Total | 156 | 158 | 158 | 161 | 162 | 161 | 161 |

**Appendix Table 4.** Ward names by council that had a high, moderate, low, and very low malaria test positivity rates in under-fives and ≥5 years from 2017 to 2023, Kagera region

| Council | Under-fives | | | | ≥ 5 years | | | |
| --- | --- | --- | --- | --- | --- | --- | --- | --- |
|  | High | Moderate | Low | Very low | High | Moderate | Low | Very low |
| Biharamulo DC* | Nyarubungo,  Runazi,Nyamigogo,Nyakahura,Nyabusozi, Nemba,Kabindi,Lusahunga,  Biharamulo Mjini,Kalenge, Kaniha,  Nyamahanga,Nyantakara | Biharamulo  Mjini,  Lusahunga,  Kalenge, Kaniha,  Nyamahanga,  Nyantakara | - | - | Biharamulo  Mjini, Kabindi,  Kaniha, Kalenge,  Nyakahura,  Nyamahanga,  Nyamigogo, Runazi, Nemba,  Lusahunga, Nyabusozi,Nyantakara,  Nyarubungo | Kalenge,  Lusahunga  ,Nemba,Nyabusozi,  Nyantakara,  Nyarubungo |  | - |
| Bukoba DC | Kibirizi, Kikomero, Rubale, Rukoma, Butelankuzi,  Butulage,  Izimbya,  Kaibanja,  Kyamulaile,Ruhunga | Buhendangabo,Bujugo,  Butelankuzi,Butulage,  Ibwera,Izimbya,Kaagya,Kaibanja,Kanyangereko,Karabagaine,Kasharu,Katoma,Katoro,Kemondo,Kishanje,Kyamulaile,Kishogo,Maruku,Mikoni,Nshambya,Nyakato,Nyakibimbili,Rubafu,Ruhunga |  |  | Kibirizi,Kikomero, Rubale, Rukoma,Buhendangabo, Bujugo,Butelankuzi,Butulage, Ibwera,  Kaagya  , Kaibanja,  Kishogo, Kyamulaile,  Nyakibimbili,Rubafu,  Ruhunga, | Buhendangabo, Bujugo,  Ibwera,Izimbya, Kaagya, Kaibanja, Kanyangereko,Bujugo,  Karabagaine, Kasharu,Katoma,Katoro,Kemondo,Kishanje, Kishogo,  Mikoni,Ruhunga,Nshambya,Nyakato,Nyakibimbili,Rubafu,Ibwera,Kyamulaile,Buhendangabo,Butelankuzi,Butulage, | Kasharu,Katoma,MarukU,  Nshambya |  |
| Bukoba MC^†^ | - |  | Bakoba,Bilele,Buhemba,Hamugembe,Ijuganyondo,Kahororo,Kitendaguro,Nyanga,Rwamishenye | Kashai,  Bakoba,Bilele,Buhemba,Hamugembe,Ijuganyondo,Miembeni |  | Buhemba,  Kahororo,  Nyanga | Bilele,Bakoba,  Buhemba,Hamugembe,Ijuganyondo,  Kahororo,Kashai,Kitendaguro,Miembeni,Nyanga,Rwamishenye | Bilele  ,Kashai,  Miembeni |
| Karagwe DC | Bugene, Bweranyange, Kihanga, Rugera,Ihembe,Kibondo,Kiruruma,Nyabiyonza,Nyakabanga,Nyakakika,Rugu | Chanika,Chonyonyo,Igurwa,Ihanda,Ihembe,Kanoni,Kayanga,Kibondo,Kiruruma,Kituntu, Ndama,Nyabiyonza,Nyaishozi,Nyakabanga,Nyakahanga,Nyakakika,Nyakasimbi |  |  | Bugene, Bweranyange,Kihanga, Nyabiyonza, Nyakabanga, Rugera, Rugu,Chonyonyo,Ihembe,Kibondo,Kiruruma,Kituntu,Nyakabanga,Nyakakika,Nyakasimbi | Chanika,Chonyonyo,Igurwa,Ihanda,Kanoni,Kayanga,Kibondo,Kiruruma,Kituntu,Ndama,Nyaishozi,Nyakabanga,Nyakahanga,Nyakakika,Nyakasimbi |  | - |
| Kyerwa DC | Kibingo, Rukuraijo,Kimuli,Kyerwa,Nkwenda,Nyakatuntu,Rwabwere,Songambele | Bugomora,Businde,Isingiro,Kaisho,Kamuli,Kibale,Kikukuru,Kimuli,Kyerwa,Mabira,Murongo,Nkwenda,Nyakatuntu,Rwabwere,Songambele |  |  | Rukuraijo,Kibale,Kibingo,Kyerwa,Mabira,Nyakatuntu,Rwabwere,Songambele | Bugomora,Businde,Isingiro,Kaisho,Kamuli,Kibale,Kibingo,Kikukuru,Kimuli,Mabira,Nkwenda,Murongo,Nyakatuntu,Rwabwere,Songambele | Isingiro  ,Kaisho | - |
| Missenyi DC | Kakunyu,Mutukula,Nsunga, | Bugandika,Bugorora,Buyango,Kakunyu,Kassambya,Kilimilil,Kitobo,Kyaka,Mabale,Minziro,Mushasha,Mutukula,Nsunga, | Bwanjai,Gera,Ishozi,Ishunju,Kanyigo,Kashenye,Ruzinga | - | Bugorora,Buyango,Ishozi,Ishunju,Kakunyu,Kanyigo,Kashenye,Kitobo,Kyaka,Mabale,Minziro,Mutukula,Nsunga | Bugandika,Bugorora,Buyango,Gera,Bwanjai,Ishozi,Ishunju,Kanyigo,Kashenye,Kassambya,Kilimilil,Kitobo,Kyaka,Mabale,Minziro,Mushasha,Nsunga,Ruzinga. | Bugandika, Bwanjai |  |
| Muleba DC | Bisheke, Bulyakashaju,Bumbire,Kabirizi,Karambi,Kasharunga, Kibanga,Kyebitembe,Mubunda,Mushabago, Ngenge, Ruhanga,Biirabo,Buganguzi,Gwanseli,Ikuza,Kashasha,Katoke,Kerebe,Kimwani,Mafumbo,Mayondwe,Mazinga,Nshamba,Nyakabango. | Biirabo,Buganguzi,Goziba,  Ikuza,Izigo,Kagoma,Kamachumu,Kashasha,Kishanda,Mayondwe,Mazinga,Muleba,Nshamba,Nyakabango |  |  | Bisheke,Bulyakashaju, Biirabo,Kabirizi,Karambi,Kasharunga, Kerebe,  Kimwani, Kyebitembe,Mushabago, Ngenge,  Ruhanga,Buganguzi,Bumbire,Gwanseli,Ikuza,Kagoma,Katoke,Kibanga,Mafumbo,Mayondwe,Mubunda,Nyakabango | Goziba,Gwanseli,Ikuza,Katoke,Izigo,Kagoma,Kamachumu,Kashasha,Kishanda,Mayondwe,Mazinga,Mubunda,Muhutwe,Muleba,Nshamba,Nyakabango,Biirabo,Buganguzi,Bumbire,Kibanga |  | - |
| Ngara DC | Bugarama, Bukiriro,Keza,Muganza,Murusagamba,Kabanga,Kanazi,Kasulo,Kibimba,Kibogora,Kirushya,Mabawe,Mbuba,Mugoma,Murukulazo,Ntobeye,Nyakisasa,Nyamiyaga,Rulenge,Rusumo. | Ngara Mjini,Kabanga,Kanazi,Kasulo,Kibimba,Kirushya,Mabawe,Mbuba,  Mugoma,Murukulazo,Ntobeye,Nyakisasa,Nyamiyaga,Rulenge,Rusumo. |  |  | Keza,Bugarama,Bukiriro,  Kabanga,Muganza,Mugoma,Murusagamba,Ntobeye,Kanazi,Kasulo,Kibimba,Kibogora,Kirushya,Mabawe,Mbuba,Murukulazo,Nyakisasa,Nyamiyaga,Rulenge,Rusumo | Mabawe,  Ngara Mjini |  | - |

*DC, district council

^†^MC, municipal council

**Appendix Table 5.** Number of wards by year in each stratum of malaria prevalence in pregnant women in Kagera region from 2017 to 2023

| Strata/Year | 2017 | 2018 | 2019 | 2020 | 2021 | 2022 | 2023 |
| --- | --- | --- | --- | --- | --- | --- | --- |
| High | 5 | 3 | 3 | 3 | - | 1 | - |
| Moderate | 116 | 82 | 81 | 96 | 85 | 77 | 91 |
| Low | 22 | 54 | 55 | 48 | 57 | 68 | 52 |
| Very low | 8 | 13 | 13 | 12 | 14 | 13 | 16 |
| Total | 151 | 152 | 152 | 159 | 156 | 159 | 159 |

**Appendix Table 6**. Ward names by council that had a high, moderate, low, and very low malaria prevalence in pregnant women from 2017 to 2023, Kagera region

| Council | High | Moderate | Low | Very low |
| --- | --- | --- | --- | --- |
| Biharamulo DC^*^ | Nyamigogo, Nyamahanga, Nemba | Biharamulo Mjini,Kabindi,  Kalenge, Kaniha,Lusahunga,  Nemba,Runazi,Nyabusozi,  Nyakahura,Nyamahanga,Nyamigogo,Nyantakara,Nyarubungo | Kabindi |  |
| Bukoba DC | Kibirizi | Butelankuzi,Butulage,Ibwera,Izimbya,Kaibanja,Kasharu,Katoro,Kibirizi,Kikomero,Kyamulaile,Nyakibimbili,Rubafu,Rubale,Ruhunga,  Rukoma | Buhendangabo,Bujugo,Kishogo,Nyakato,Ibwera,Kaagya,Kaibanja,Kanyangereko,Karabagaine,Kasharu,Katoma,Katoro,Kemondo,Kishanje, Maruku,Mikoni | Kaagya,  Kanyangereko,Nyakato,Karabagaine,Katoma, Kishogo,  Mikoni |
| Bukoba MC^†^ |  |  | Bakoba,Bilele,Buhemba,Ijuganyondo,Kahororo,Kashai,Kitendaguro,Miembeni,Nshambya,Rwamishenye | Bilele,Kashai,  Kitendaguro,  Miembeni,Nyanga |
| Karagwe DC | Bugene, Bweranyange | Bugene,Bweranyange,Chonyonyo,Ihanda,Ihembe,Kanoni,Kayanga,Kihanga,Kiruruma,Ndama,Nyabiyonza,Nyaishozi,Nyakabanga,Nyakahanga,Nyakakika,Nyakasimbi,Rugera,Rugu | Chanika,Igurwa, Ihanda,  Ihembe,Kanoni,Kayanga,Kibondo,Kiruruma,Kituntu,Nyabiyonza,Nyaishozi |  |
| Kyerwa DC | Nkwenda | Isingiro,Kibingo,Kyerwa,Mabira,Nkwenda,Nyakatuntu,Rukuraijo,Rwabwere,Songambele | Bugomora,Businde,Isingiro,Kaisho,Kamuli,Kibale,Kibingo,Kikukuru,Kimuli,Kyerwa,Mabira,Murongo,Nyakatuntu | Bugomora |
| Missenyi DC |  | Bugorora,Buyango,Ishozi,  Ishunju,Kakunyu,Kassambya,Kilimilil,Kyaka,Mabale,Minziro,Mushasha,Mutukula,Nsunga | Bugandika,Bugorora,  Buyango,Bwanjai,Gera,  Ishozi,Kakunyu,Kanyigo,  Kashenye,Kitobo,Ruzinga | Bugandika,  Bwanjai,  Kanyigo,  Ruzinga |
| Muleba DC | Mubunda, Bisheke, Kibanga, Ruhanga | Biirabo,Bisheke,Buganguzi,  Bulyakashaju,Gwanseli,Kabirizi,Kagoma,Kamachumu,Karambi,Kasharunga,Katoke,Kibanga,Kimwani,Kishanda,Kyebitembe,Mafumbo,Mazinga,Mubunda,Mushabago,Ngenge,Nshamba,  Nyakabango,Ruhanga | Mayondwe,Bumbire,  Goziba,Ikuza,Izigo,Kabirizi,Kagoma,Kamachumu,Kashasha,Kerebe,Mayondwe,  Muhutwe,Muleba,  Nshamba |  |
| Ngara DC |  | Bugarama,,Bukiriro,Keza,  Kabanga,Kanazi,Kasulo,Kibimba,Kibogora,Mabawe,Mbuba,Muganza,Murukulazo,Murusagamba,Ngara Mjini,Ntobeye,Nyamiyaga,  Rulenge,Rusumo | Kirushya,Mugoma,  Murukulazo,Murusagamba,  Ntobeye,Nyakisasa | Kirushya |

*DC, district council

^†^MC, municipal council

**Appendix E. School Malaria Parasitological Survey (SMPS)**

School malaria parasitological surveys (SMPS) were first conducted in 31 schools (2799 children) across the eight councils in 2015 (*13*) and this was increased to 34 schools (3516 children) in 2017 to accommodate the expansion of administrative boundaries in 2016 and in 2019, 33 schools (3425 children) were selected. In 2021, a total of 34 schools (3325 children) were selected for the survey. In total, 40 different schools from 37 selected wards of Kagera region were sampled.

**Appendix Table 7.** Number of wards by year in each stratum of malaria prevalence in school children in Kagera region from 2017 to 2023

| Strata/Year | 2015 | 2017 | 2019 | 2021 |
| --- | --- | --- | --- | --- |
| High | 17 | 19 | 13 | 8 |
| Moderate | 9 | 9 | 12 | 16 |
| Low | 2 | 5 | 6 | 8 |
| Very low | 3 | 1 | 2 | 2 |
| Total | 31 | 34 | 33 | 34 |

**Appendix Table 8**. Ward names by council that had a high, moderate, low, and very low malaria prevalence in school children from 2017 to 2023, Kagera region

| Council | High | Moderate | Low | Very low |
| --- | --- | --- | --- | --- |
| Biharamulo DC* | Kaniha, Lusahunga, Nyamigogo,  Biharamulo Mjini | Biharamulo Mjini, Kaniha,  Lusahunga |  |  |
| Bukoba DC | Mugajwale, Rukoma |  | Katoma,  Maruku,  Rubafu |  |
| Bukoba MC^†^ |  |  | Kitendaguro,Bilele,Buhemba | Bilele, Buhemba, |
| Karagwe DC | Rugera,  Bweranyange, | Bweranyange,Chonyonyo,Rugu,Rugera | Chonyonyo |  |
| Kyerwa DC | Rukuraijo | Kibingo,Kikukuru,Mabira | Kibingo, |  |
| Missenyi DC | Mutukula,  Mushasha | Bugandika,  Mushasha, Mabale | Bugandika |  |
| Muleba DC | Kimwani,  Bisheke,  Nyakatanga | Bisheke,  Katoke,Kishanda,Kyebitembe,  Nyakatanga,  Kimwani | Kamachumu,Muleba | Kamachumu |
| Ngara DC | Bugarama, Nyamiyaga,  Murukulazo,  Ntobeye,Rulenge | Ntobeye,  Rulenge |  |  |

*DC, district council

^†^MC, municipal council

**Appendix Table 9.** Overall malaria burden at ward level in Kagera region by years and study groups from 2015 to 2023

| Year/  Group | ANC* | | SMPS^†^ | | Under-fives | | ≥ 5 years | |
| --- | --- | --- | --- | --- | --- | --- | --- | --- |
|  | Prev.  (%) | Rate  (%) | Prev.  (%) | Rate  (%) | TPR^‡^  (%) | Rate  (%) | TPR  (%) | Rate  (%) |
| 2015 |  |  | 31.1 | - |  |  |  |  |
| 2016 |  |  |  |  |  |  |  |  |
| 2017 | 12.3 | - | 29.2 | -6.1 | 40.8 | - | 39.7 | - |
| 2018 | 9.1 | -26.0 |  |  | 28.2 | -30.9 | 28.8 | -27.5 |
| 2019 | 8.8 | -3.3 | 27.2 | -6.8 | 36.6 | +29.8 | 35.4 | +22.9 |
| 2020 | 9.1 | +3.4 |  |  | 35.6 | -2.7 | 36.4 | +2.8 |
| 2021 | 8.4 | -4.5 | 17.4 | -36.0 | 30.1 | -15.4 | 31.6 | -14.6 |
| 2022 | 6.8 | -1.9 |  |  | 29.8 | -0.9 | 31.1 | -1.6 |
| 2023 | 7.8 | +14.7 |  |  | 32.6 | +9.4 | 34.7 | +11.6 |
| Total | 10.0 | -36.6 | 26.1 | -44.1 | 33.1 | -20.1 | 33.7 | -12.6 |

“Malaria burden is different in each year (all p < 0.001) when comparing to reference 2017)”

*ANC = Antenatal care; ^†^SMPS = School malaria parasitological survey; ^‡^TPR = Test positivity rate.

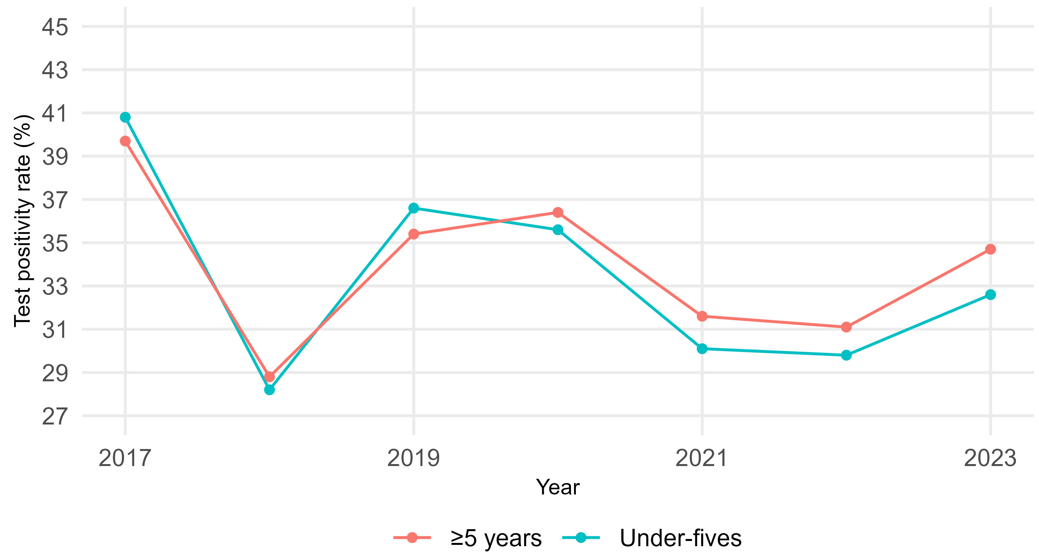

**Appendix Figure 2**. Comparison of malaria test positivity rates in Kagera region from 2017 to 2023 based on the data from health facilities.

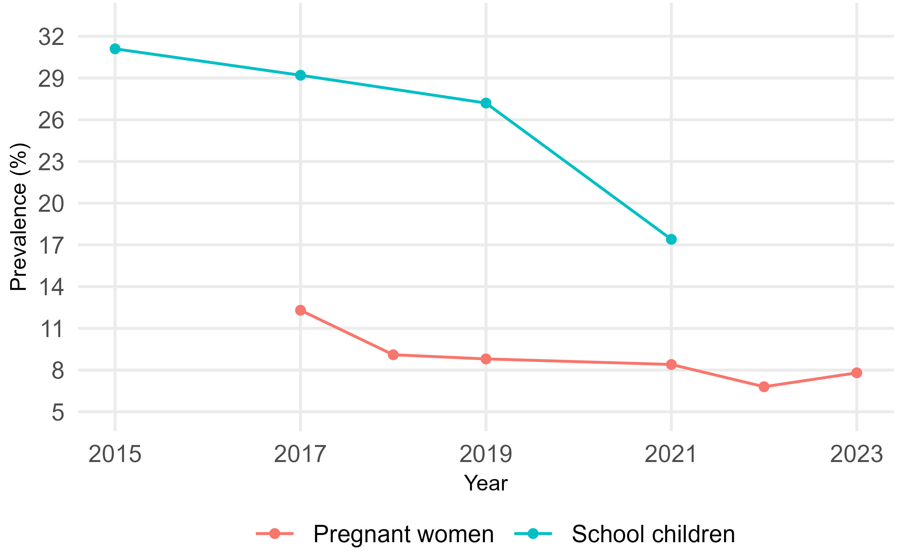

**Appendix Figure 3**. Comparison of malaria prevalence in Kagera region from 2017 to 2023 based on the data from pregnant women and school children.

**Appendix Table 10.** Names of hotspots of malaria burden at ward level in Kagera region at 95% and 99% confidence levels from 2017 to 2023

| Group | Year | Council | 95% CL^‡^ | 99% CL |
| --- | --- | --- | --- | --- |
| ANC*  ANC | 2017 | Biharamulo DC^†^ | Kaniha,Biharamulo Mjini, Nyabusozi, Kalenge, Nyarubungo,Kabindi,Nyantakara,  Nyamigogo, Nyamahanga, Lusahunga | Biharamulo Mjini, Lusahunga, Kabindi, Nyarubungo |
|  |  | Muleba DC | Nyakabango,Kimwani,Mubunda, Kibanga,  Kamachumu, Kyebitembe, Bulyakashaju | Bulyakashaju |
|  |  | Kyerwa DC | Rwabwere, Songambele,Nkwenda | Songambele |
|  | 2018 | Biharamulo DC | Nyantakara, Biharamulo Mjini, Nyarubungo, Kabindi,Lusahunga,Nyamahanga,Nyamigogo, Runazi | Nyarubungo, Kabindi, Biharamulo Mjini, Lusahunga, Nyamahanga,  Nyamigogo, Runazi |
|  |  | Muleba DC | Gwanseli, Kimwani, Kyebitembe,  Mubunda,Bisheke,Kibanga,Nyabusozi,Nyakabango,Karambi,Kasharunga | Bisheke,Kyebitembe,Kasharunga, Kibanga,Nyabusozi,Mubunda, Nyakabango, Kimwani,Karambi |
|  |  | Karagwe DC | Rugu | Rugu |
|  | 2019 | Biharamulo DC | Nyantakara, Nyamigogo, Biharamulo Mjini, Nyamahanga, Nyarubungo,Runazi, Lusahunga, Kalenge, Kabindi | Nyantakara, Nyamigogo, Nyarubungo,Runazi, Lusahunga,  Kabindi |
|  |  | Muleba DC | Karambi, Kimwani, Nyabusozi, Kibanga, Mubunda, Kasharunga, Ngenge, Kyebitembe, Nyakabango, Bisheke | Nyabusozi, Kibanga, Mubunda,  Kasharunga, Kyebitembe, Bisheke |
|  |  | Karagwe DC | Rugu, Kayanga, Rugera | Kayanga, Rugera |
|  |  | Bukoba DC | Ruhunga, Kibirizi, Butulage,  Bugene,Rubale, Rukoma | Kibirizi, Butulage, Rukoma |
|  | 2020 | Muleba DC | Mushabago, Ngenge, Mubunda, Bisheke, Kasharunga, Kibanga | Ngenge, Mubunda, Bisheke, Kasharunga, Kibanga |
|  |  | Biharamulo DC | Kabindi, Nyarubungo, Nyabusozi, Runazi, Nyantakara, Nemba, Lusahunga, Kaniha | Nyarubungo, Nyabusozi, Nyantakara, Nemba, Lusahunga, Runazi, Kaniha |
|  |  | Karagwe DC | Rugu, Rugera | Rugera |
|  |  | Missenyi DC | Kilimilil, Mabale |  |
|  |  | Bukoba DC | Kyamulaile, Butulage, Rubale, Kibirizi, Rukoma, Ruhunga | Kyamulaile, Butulage, Rubale, Kibirizi, Rukoma, Ruhunga |
|  | 2021 | Biharamulo DC | Nyamahanga, Nyamigogo, Nyantakara, Nyakahura, Runazi, Lusahunga, Nemba, Nyabusozi,Nyarubungo, Kabindi | Runazi, Lusahunga, Nemba, Nyabusozi,Nyarubungo, Kabindi |
|  |  | Muleba DC | Ngenge, Kimwani, Kyebitembe, Kasharunga, Mubunda, Bisheke, Kibanga | Kasharunga, Mubunda, Bisheke, Kibanga |
|  |  | Ngara DC | Murusagamba, Keza, Rulenge, Bukiriro, Muganza, Nyakisasa | Muganza, Nyakisasa |
|  |  | Karagwe DC | Rugu, Rugera | Rugera |
|  |  | Bukoba DC | Kyamulaile, Izimbya, Ruhunga, Rubale,Rukoma, Kibirizi, Butulage | Ruhunga, Rubale,  Rukoma, Kibirizi, Butulage |
|  | 2022 | Biharamulo DC | Nyantakara, Lusahunga | Lusahunga |
|  |  | Ngara DC | Murusagamba, Keza, Nyakisasa,Bugarama, Muganza, Rulenge, Bukiriro | Muganza, Rulenge, Bukiriro |
|  |  | Karagwe DC | Rugu, Rugera |  |
|  |  | Muleba DC | Ngenge, Mubunda, Bisheke, Kasharunga, Kibanga | Mubunda, Bisheke, Kasharunga, Kibanga |
|  |  | Bukoba DC | Izimbya, Butulage, Rukoma, Kibirizi, Rubale | Butulage, Rukoma, Kibirizi, Rubale |
|  | 2023 | Muleba DC | Kasharunga, Kibanga, Mubunda, Bisheke | Kibanga, Mubunda, Bisheke |
|  |  | Ngara DC | Bukiriro, Rulenge, Nyakisasa | Nyakisasa |
|  |  | Biharamulo DC | Nyabusozi, Kabindi, Lusahunga, Nemba,Runazi,Nyantakara | Lusahunga, Nemba |
|  |  | Karagwe DC | Rugera, Rugu | Rugu |
|  |  | Bukoba DC | Ruhunga, Butulage, Rubale,  Rukoma, Izimbya, Kibirizi | Butulage, Rubale,Rukoma, Izimbya, Kibirizi |
| Under-  fives | 2017 | Bukoba DC | Kikomero | Kikomero |
|  |  | Biharamulo DC | Biharamulo Mjini, Lusahunga, Nyamahanga, Nyarubungo | Nyarubungo |
|  |  | Muleba DC | Kamachumu, Ngenge, Katoke, Nyakabango, Karambi, Biirabo, Kimwani, Kibanga, Bulyakashaju,Kyebitembe, Bisheke, Mubunda Mushabango, Kabirizi, Kasharunga, | Kimwani, Kibanga, Bulyakashaju,  Kyebitembe, Bisheke, Mushabango, Kabirizi, Kasharunga  Mubunda |
|  |  | Ngara DC | Bugarama, Bukiriro, Muganza |  |
|  |  | Kyerwa DC | Rukuraijo |  |
|  | 2018 | Biharamulo DC | Biharamulo Mjini,Nyamigogo,  Kalenge, Runazi, Lusahunga,  Nyantakara,Nyamahanga,  Nyarubungo, Nyabusozi, Kabindi | Runazi, Nyantakara, Nyamahanga, Lusahunga, Nyarubungo, Nyabusozi, Kabindi |
|  |  | Muleba DC | Bulyakashaju, Biirabo, Mubunda,  Kimwani,Kasharunga, Kyebitembe,  Kibanga, ,Bisheke,Karambi,Kabirizi,  Ngenge, Nyakabango, Mushabango | Mubunda, Kasharunga, ,Bisheke,  Kyebitembe, Kibanga, Karambi,  Kabirizi, Ngenge, Nyakabango,  Kimwani, Mushabango |
|  |  | Karagwe DC | Rugu | Rugu |
|  |  | Bukoba DC | Rukoma, Kikomero | Kikomero |
|  | 2019 | Biharamulo DC | Nyakahura, Lusahunga,Nyarubungo | Lusahunga,Nyarubungo |
|  |  | Muleba DC | Bisheke, Nyakabango, Kimwani,  Mubunda, Kasharunga, Kyebitembe  ,Mushabango, Ngenge, Karambi | Mubunda, Kasharunga,  Mushabango, Ngenge, Karambi,  Kyebitembe |
|  |  | Karagwe DC | Rugu, Rugera |  |
|  |  | Ngara DC | Mabawe, Kanazi, Kabanga, , Keza  Muganza, Bukiriro, Bugarama,  Rulenge, Mbuba, Murusagamba, Nyakisasa | Muganza, Bukiriro, Bugarama,  Rulenge, Mbuba, Murusagamba,  Nyakisasa, Keza |
|  |  | Bukoba DC | Ruhunga, Kikomero, Rubale,  Butulage,Rukoma, Kibirizi | Kikomero, Butulage,Rukoma,  Rubale, Kibirizi |
|  | 2020 | Ngara DC | Kabanga, Kanazi, Kibimba, Rulenge,  Mugoma,Bugarama, Bukiriro, Nyakisasa, Mbuba, Keza, Murusagamba,Muganza | Bugarama, Bukiriro, Nyakisasa,  Mbuba, Keza, Murusagamba,  Muganza, Rulenge |
|  |  | Biharamulo DC | Nyakahura | Nyakahura |
|  |  | Muleba DC | Kasharunga, Bisheke, Mubunda,  Ngenge, Mushabago | Mubunda, Ngenge, Mushabago |
|  |  | Karagwe DC | Rugera |  |
|  |  | Bukoba DC | Kyamulaile, Butelankuzi, Izimbya,  Rukoma, Rubale, Kikomero,  Ruhunga, Butulage, Kibirizi | Rukoma, Rubale, Kikomero,  Ruhunga, Butulage, Kibirizi |
|  | 2021 | Ngara DC | Ngara Mjini, Kirushya, Murukulazo, Kibimba,  Nyamiyaga, Nyakisasa, Keza, Mugoma,  Mbuba, Muganza, Bugarama, Kabanga,  Bukiriro, Rulenge, Rusumo, Kanazi,  Murusagamba, Ntobeye, Mabawe, | Nyakisasa, Keza, Mbuba,  Muganza, Bugarama, Bukiriro,  Rulenge, Rusumo, Kanazi,  Murusagamba, Ntobeye, Mabawe,  Kibimba, Mugoma, Kabanga |
|  |  | Biharamulo DC | Nyakahura | Nyakahura |
|  |  | Bukoba DC | Ruhunga, Rubale, Rukoma,  Kibirizi, Butulage, Kikomero | Rukoma, Kibirizi, Butulage,  Kikomero |
|  |  | Muleba DC | Mushabango, Ngenge | Mushabango, Ngenge |
|  | 2022 | Biharamulo DC | Nyakahura |  |
|  |  | Ngara DC | Ngara Mjini, Kirushya, Rulenge, Kibimba,  Keza,Murusagamba, Kabanga, Kanazi,,  Rusumo, Nyakisasa, Bugarama,  Muganza, Mugoma, Murukulazo, Bukiriro  Mbuba, Nyamiyaga, Ntobeye | Rusumo, Nyakisasa, Bugarama,  Muganza, Mugoma, Murukulazo,  Mbuba, Nyamiyaga, Ntobeye,  Rulenge, Kibimba, Kanazi, Bukiriro |
|  |  | Muleba DC | Kabirizi, Ngenge, Mushabago | Ngenge, Mushabago |
|  |  | Bukoba DC | Ruhunga, Rubale, Rukoma,  ,Butulage, Kikomero, Kibirizi | Rubale, ,Butulage, Kikomero, Kibirizi |
|  | 2023 | Ngara DC | Ngara Mjini, Keza, Kirushya,  Kabanga,Kibimba,Nyamiyaga,Muruk  ulazo,Mugoma,Ntobeye,Kanazi,Rus  umo, Bugarama,Nyakisasa,  Muganza, Mbuba, Bukiriro, Rulenge | Kibimba,Nyamiyaga,Murukulazo,  Mugoma,Ntobeye,Kanazi,Rusumo,  Bugarama,Nyakisasa,  Muganza, Mbuba, Bukiriro,  Rulenge |
|  |  | Muleba DC | Ngenge, Mushabago | Ngenge, Mushabago |
|  |  | Bukoba DC | Ruhunga, Rubale, ,Rukoma,  Kikomero, Kibirizi,Butulage | Rubale, ,Rukoma, Kikomero,  Kibirizi,Butulage |
| ≥ 5 years  ≥ 5 years | 2017 | Biharamulo DC | Nyarubungo | Nyarubungo |
|  |  | Kyerwa DC | Nkwenda, Rukuraijo,Kikukuru | Rukuraijo |
|  |  | Muleba DC | Biirabo, Buganguzi, Bisheke, Kasharunga,Kabirizi,Bulyakashaju, Ngenge, Mushabago, Nyakabango ,Kamachumu,Kimwani,Katoke,Mubunda,Kyebitembe,Kibanga, Karambi | Bisheke, Bulyakashaju, Ngenge, Kasharunga,Kabirizi, Mushabago, Kyebitembe,Kibanga, Mubunda, Katoke |
|  |  | Bukoba DC | Kikomero | Kikomero |
|  | 2018 | Biharamulo DC | Lusahunga, Nyamahanga, Kalenge, Nyantakara, Nyarubungo, Runazi Nyamigogo, Nyabusozi, Kabindi | Lusahunga, Nyarubungo, Nyamigogo, Nyabusozi, ,Kabindi, Runazi |
|  |  | Muleba DC | Kamachumu,Biirabo, Bulyakashaju, Bisheke, Kibanga, Ngenge, Nyakabango, Kabirizi, Karambi, Kasharunga, Kimwani, Kyebitembe, Mushabago,Mubunda | Bisheke, Nyakabango, Kabirizi, Karambi, Kasharunga, Kimwani, Kyebitembe, Mushabago,Mubunda |
|  |  | Karagwe DC | Rugu | Rugu |
|  |  | Bukoba DC | Rukoma, Kikomero | Kikomero |
|  | 2019 | Ngara DC | Bugarama, Nyakisasa, Keza, Rulenge | Rulenge |
|  |  | Muleba DC | Karambi, Ngenge, Bisheke, Mubunda, Kimwani, Nyakabango, Bulyakashaju, Kasharunga,Mushabago, Kabirizi,Kyebitembe | Kasharunga,  ,Mushabago, Kabirizi, Kyebitembe, |
|  |  | Ngara DC | Kirushya, Bukiriro, Mbuba,  ,Murusagamba, Muganza |  |
|  |  | Biharamulo DC | Nyakahura, Nyarubungo | Nyarubungo |
|  |  | Bukoba DC | Kibirizi, Rubale, Butulage, Rukoma, Kikomero | Rubale, Butulage, Rukoma, Kikomero |
|  | 2020 | Muleba DC | Mubunda, Bisheke, Kabirizi, Bulyakashaju, Ngenge, Kasharunga, Mushabago | Kasharunga, Mushabago |
|  |  | Ngara DC | Bugarama, Kirushya, Rusumo, Bukiriro, Mugoma, Mbuba, Murusagamba,Muganza, ,Keza, Nyakisasa, Rulenge | Mbuba, Murusagamba,Muganza, ,Keza, Nyakisasa, Rulenge |
|  |  | Biharamulo DC | Nyakahura |  |
|  |  | Bukoba DC | Butelankuzi, Izimbya, Ruhunga, Kibirizi, Butulage, Rukoma,Kikomero, Rubale | Butulage, Rukoma,Kikomero, Rubale |
|  | 2021 | Ngara DC | Ngara Mjini, Mabawe, Kabanga, Kasulo, Bugarama, Murusagamba, Rusumo, Murukulazo ,Muganza, Mugoma ,Mbuba, Nyakisasa, Kirushya, Nyamiyaga, Kibimba, Keza,Bukiriro, Ntobeye, Kanazi, Rulenge | Bugarama, Murusagamba,Rusumo,  Murukulazo,Muganza, Mugoma ,Mbuba, Nyakisasa, Kirushya, Nyamiyaga, Kibimba, Keza,Bukiriro,  Ntobeye, Kanazi, Rulenge |
|  |  | Muleba DC | Ngenge, Mushabango | Ngenge, Mushabango |
|  |  | Bukoba DC | Rubale, Butulage, Kibirizi, Kikomero, Rukoma | Butulage, Kibirizi, Kikomero, Rukoma |
|  |  | Biharamulo DC | Nyakahura | Nyakahura |
|  | 2022 | Biharamulo DC | Nyakahura |  |
|  |  | Ngara DC | Ngara Mjini, Kibimba, Murusagamba, Bugarama, Rusumo, Keza, Kanazi, Ntobeye. Rulenge, Bukiriro, Nyamiyaga, Kirushya Nyakisasa, Mugoma,Mbuba,Murukulazo, Muganza | Murusagamba, Bugarama, Rusumo, Keza, Kanazi, Ntobeye. Rulenge, Bukiriro, Nyamiyaga, Kirushya Nyakisasa, Muganza, Mugoma,Mbuba,Murukulazo |
|  |  | Muleba DC | Kabirizi, Ngenge, Mushabago | Ngenge, Mushabago |
|  |  | Bukoba DC | Ruhunga, Butulage, Rubale, Kibirizi, Rukoma, Kikomero | Butulage, Rubale, Kibirizi, Rukoma,  Kikomero |
|  | 2023 | Ngara DC | Kanazi, Murusagamba, Kibimba, Nyamiyaga, Mugoma, Mbuba, Ntobeye, Rulenge,Rusumo,Kirushya,Keza,Bugarama, Bukiriro,Muganza,Nyakisasa, Murukulazo, | Nyamiyaga, Mugoma, Rusumo,Kirushya,Keza,Bugarama, Bukiriro,Muganza,Nyakisasa, Murukulazo, Mbuba, Ntobeye, Rulenge |
|  |  | Biharamulo DC | Nyakahura |  |
|  |  | Muleba DC | Mushabago, Bulyakashaju |  |
|  |  | Bukoba DC | Ruhunga, Butulage, Rukoma, Kikomero, Kibirizi, Rubale | Butulage, Rukoma, Kikomero, Kibirizi, Rubale |

*ANC = Antenatal Care, ^†^DC, district council, ^‡^CL= Confidence Level

**Appendix Table 11.** Names of coldspots of malaria burden at ward level in Kagera region at 95% and 99% confidence levels from 2017 to 2023

| Group | Year | Council | 95% CL | 99% CL |
| --- | --- | --- | --- | --- |
| ANC  ANC | 2017 | Bukoba DC | Kaagya, Karabagaine, Kishanje, Kanyangereko,Buhendangabo,Katoma, Maruku, Bujugo, Kemondo, Nyakato, | Maruku, Bujugo, Kemondo, Nyakato, |
|  |  | Bukoba MC | Kashai,Bilele,Nshambya,  Kitendaguro, Ijuganyondo,Buhemba, Bakoba, Miembeni, Kahororo | Bakoba, Miembeni, Kahororo |
|  |  | Missenyi DC | Bwanjai, Gera, Bugandika, Ishunju, Kanyigo, |  |
|  | 2018 | Bukoba MC | Kahororo, Nshambya, Miembeni, Kashai, |  |
|  |  | Bukoba DC | Bujugo, Nyakato, Maruku, Karabagaine, Kemondo, Kasharu |  |
|  |  | Missenyi DC | Kanyigo, Bugandika, Bugorora, Buyango |  |
|  | 2019 | Bukoba DC | Maruku,Karabagaine,Nyakato,Bujugo,Buhendangabo,Kemondo,Kasharu,  Kaagya | Bujugo |
|  |  | Bukoba MC | Kahororo,Bilele,Nshambya,Bakoba,  Kashai, Miembeni |  |
|  |  | Missenyi DC | Bugandika, Bwanjai, Gera, Kanyigo, Ishozi, Buyango |  |
|  |  | Ngara DC | Ntobeye |  |
|  | 202  2020 | Bukoba MC | Nshambya, Miembeni, Kashai, Bilele, Kahororo, Bakoba, Buhemba,Nyanga |  |
|  |  | Bukoba DC | Nyakato, Buhendangabo,Bujugo, Kaagya, Maruku, Kemondo, Katoma, Kishanje, Karabagaine | Karabagaine |
|  |  | Missenyi DC | Ishozi, Bwanjai, Bugandika, Kanyigo, Gera | Kanyigo, Gera |
|  | 2021 | Kyerwa DC | Kibale |  |
|  |  | Bukoba MC | Nshambya,Kahororo,Kashai,Nyanga, Miembeni,Bakoba, Bilele, Ijuganyondo |  |
|  |  | Bukoba DC | Bujugo, Katoma, Buhendangabo, Kemondo,Kaagya,Nyakato,Maruku,  Karabagaine | Maruku,Karabagaine |
|  |  | Missenyi DC | Gera, Ishozi,Bwanjai, Bugandika, Kanyigo |  |
|  | 2022 | Bukoba MC | Bakoba, Nshambya |  |
|  |  | Bukoba DC | Karabagaine,Katoma,Kasharu,  Nyakato,Bujugo,Maruku |  |
|  |  | Missenyi DC | Kanyigo,Gera, Bugandika |  |
|  | 2023 | Bukoba MC | Kashai,Nyanga,Buhemba,Nshambya, Kahororo |  |
|  |  | Bukoba DC | Kemondo,Karabagaine,Maruku, ,Nyakato. Katoma |  |
|  |  | Missenyi DC | Gera |  |
|  |  | Kyerwa DC | Kamuli, Nyakatuntu,Rutunguru, Kibale, Mabira |  |
| Under-fives | 2017 | Bukoba MC | Buhemba,Bilele,Miembeni,Bakoba,  Kashai,Nshambya,Ijuganyondo Kahororo,Kitendaguro | Bilele,Miembeni,Bakoba,Kashai,  Nshambya, Kahororo,Kitendaguro,  Ijuganyondo |
|  |  | Bukoba DC | Kasharu, Kanyangereko, Kaagya,  Kishogo, Maruku, Katoro,  Bujugo,Katoma,Karabagaine,  Kishanje, Kaibanja, Buhendangabo | Maruku, Bujugo, Karabagaine, Katoro, Katoma, Kishanje, Kaibanja, Buhendangabo |
|  |  | Missenyi DC | Kashenye, Ishunju, Nsunga. Bugorora, Bugandika, Gera, Mushasha,Kitobo ,Buyango, Minziro, Bwanjai,Ishozi, Kanyigo,Kassambya, Kyaka, Ruzinga | Bugorora,Bugandika,Gera,Buyango, Minziro, Bwanjai,Ishozi,  Kanyigo,Kassambya,Mushasha,  Kitobo, Kyaka, Ruzinga |
|  | 2018 | Bukoba MC | Kahororo,Miembeni,Kashai, Bakoba, Kitendaguro,Ijuganyondo, Bilele, Rwamishenye, Nshambya, Hamugembe | Bilele, Nshambya, Hamugembe |
|  |  | Bukoba DC | Kishanje,Kaagya,Kasharu, Buhendangabo,Katoro,Kaibanja, Katoma,Kishogo,Kanyangereko, Nyakibimbili,Bujugo,Maruku, Karabagaine | Bujugo,Maruku, Karabagaine |
|  |  | Missenyi DC | Kyaka, Kashenye, Nsunga,  Mushasha,Ruzinga,Kitobo, Ishunju, Mutukula, Bugorora, Bugandika, Bwanjai, Kassambya,  Kanyigo,Minziro,Bunyago,Gera,Ishozi, | Bugorora, Bugandika,  Kanyigo,Minziro, Bunyago,Gera,Ishozi, Bwanjai, Kassambya |
|  | 2019 | Bukoba MC | Rwamishenye,Buhemba,Nshambya,  Bilele,Bakoba,Kahororo,Miembeni,  Kashai,Hamugembe,Kitendaguro,  Ijuganyondo | Nshambya,Bilele,Bakoba, Kahororo,Miembeni,Kashai, Hamugembe,Kitendaguro,  Ijuganyondo |
|  |  | Bukoba DC | Kasharu,Kemondo,Kishogo, Nyakibimbili,Maruku,Bujugo, Kishanje,  Karabagaine, Kaagya,Katoma,  Buhendangabo, Kanyangereko, | Maruku,Bujugo,Karabagaine, Kaagya,Buhendangabo, Katoma, Kanyangereko, Kishanje |
|  |  | Missenyi DC | Bugorora, Kitobo, Ruzinga,Kashenye, Minziro,Gera,Ishozi,Ishunju, Bugandika,Kanyigo,Bwanjai,Buyango | Gera, Ishozi, Bugandika,Kanyigo, Bwanjai, Buyango |
|  | 2020 | Bukoba MC | Rwamishenye,Kahororo,Bilele,  Nshambya,Bakoba,Miembeni,  Hamugembe, Kashai, Kitendaguro, Ijuganyondo, Nyanga, Buhemba | Bilele,Nshambya,Bakoba, Miembeni,Hamugembe, Kashai, Kitendaguro, Ijuganyondo, Buhemba |
|  |  | Bukoba DC | Kishanje,Kanyangereko,Maruku, Karabagaine,Nyakato,Katoma, Kemondo,Buhendangabo,Bujugo,  Kaagya, Maruku | Maruku, Karabagaine, Nyakato, Katoma,Bujugo,Kaagya, Buhendangabo, Kemondo, Maruku |
|  |  | Missenyi DC | Ishunju,Buyango,Ruzinga, Kitobo, Kashenye, Gera, Kanyigo, Ishozi,Bwanjai, Bugandika | Gera, Kanyigo, Ishozi,Bwanjai, Bugandika |
|  |  | Muleba DC | Mayondwe |  |
|  | 2021 | Bukoba MC | Nyanga, Bileke,Bakoba, Nshambya, Miembeni,Kashai,Kahororo, Kitendaguro,Ijuganyondo,Hamugembe | Bileke,Bakoba, Nshambya, Miembeni, Kashai, Kahororo, Kitendaguro, Ijuganyondo, Hamugembe |
|  |  | Bukoba DC | Kemondo, Kishanje,Kanyengereko, Karabagaine,Katoma, Nyakato, Buhendangabo,Kaagya, Bujugo | Karabagaine,Katoma, Nyakato, Buhendangabo,Kaagya, Bujugo |
|  |  | Missenyi DC | Bugorora, Kitobo, Kashenye,Ishunju, Bwanjai,Gera,Kanyigo,Ishozi, Bugandika, Buyango, Ruzinga | Bwanjai, Gera,Kanyigo,Ishozi, Bugandika, Buyango, Ruzinga, |
|  | 2022 | Bukoba MC | Rwamishenye,Nshambya,Kahororo,  Miembeni,Bilele,Kashai,Bakoba, Buhemba, Nyanga, Kitendaguro, Ijuganyondo | Nshambya,Kahororo,Miembeni,Bilele,Kashai,Bakoba, Buhemba, Nyanga, Kitendaguro, Ijuganyondo |
|  |  | Bukoba DC | Buhendangabo, Kanyangereko, Maruku, Karabagaine, Nyakato, Katoma, Bujugo | Maruku, Karabagaine, Nyakato, Katoma, Bujugo |
|  |  | Missenyi DC | Ishozi, Buyango,Bugorora, Kitobo, Gera, Bugandika, Bwanjai,Kanyigo | Gera, Bugandika, Bwanjai,Kanyigo |
|  | 2023 | Bukoba MC | Kitendaguro,Ijuganyondo,Kibeta, Nshambya,Miembeni,Bilele,Kahororo,  Kashai, Buhemba,Bakoba,Nyanga | Nshambya,Miembeni,Bilele,Kahororo,  Kashai, Buhemba,Bakoba,Nyanga, Kibeta |
|  |  | Bukoba DC | Nyakato,Katoma,Bujugo,Maruku,  Karabagaine, | Maruku |
|  |  | Kyerwa DC | Mabira |  |
|  |  | Karagwe | Chanika |  |
| ≥5 years | 2017 | Bukoba MC | Bilele, Bakoba,Miembeni,Kashai, Kitendaguro,Nsambya, Ijuganyondo,Kahororo | Bilele, Bakoba,Miembeni,Kashai, Kitendaguro, Nsambya, Ijuganyondo,Kahororo |
|  |  | Bukoba DC | Kishanje,Kanyengereko, Kishogo, Kaagya,Kemondo, Buhengangabo, Maruku,Bujugo,Kasharu,Kaibanja,  Karabagaine,Katoro | Maruku,Bujugo,Kasharu,Kaibanja,  Karabagaine,Katoro |
|  |  | Missenyi DC | Ruzinga,Ishunju, Nsunga,Kashenye, Bugorora,Bugandika,Mushasha,  Kanyigo,Bwanjai,Kyaka,Minziro, Kitobo, Buyango,Kassambya,Gera,Ishozi, | Bugorora,Bugandika,Mushasha,  Kanyigo,Bwanjai,Kyaka,Minziro, Buyango,Kassambya,Gera,Ishozi,  Kitobo |
|  | 2018 | Bukoba MC | Kashai, Kahororo, Kitendaguro, Ijuganyondo,Rwamishenye,Bilele,  Bakoba,Nshambya, Hamugembe, Miembeni | Bilele,Bakoba,Nshambya, Hamugembe, Miembeni |
|  |  | Bukoba DC | Kaibanja,Kishanje,Kaagya, Kasharu,  Kemondo,Buhendangabo, Kanyangereko, Maruku,Bujugo, Katoro,Karabagaine | Maruku,Bujugo, Katoro,Karabagaine |
|  |  | Missenyi DC | Mushasha, Ruzinga,Kitobo,Kyaka,  Kashenye, Ishunju, Nsunga, Mutukula, Bugorora, Bugandika, Minziro,Gera,Buyango,Kanyingo,  Ishozi,Bwanjai, Kassambya | Bugorora, Bugandika, Minziro,Gera, Buyango,Kanyingo,Ishozi,Bwanjai, Kassambya |
|  | 2019 | Bukoba MC | Buhemba, Bilele,Bakoba, Nshambya, Hamugembe,Miembeni,Kashai, Kahororo,Kitendaguro, Ijuganyondo, Rwamishenye | .Bilele,Bakoba,Nshambya, Hamugembe,Miembeni,Kashai, Kahororo,Kitendaguro, Ijuganyondo, Rwamishenye |
|  |  | Bukoba DC | Katoma,Kemondo, Kishogo, Kaagya, Kishanje,Kaibanja, Maruku, Bujugo, Karabagaine,Buhendangabo,Kasharu,Kanyangereko | Maruku, Bujugo, Karabagaine, Buhendangabo,Kasharu,  Kanyangereko |
|  |  | Missenyi DC | Kashenye, Kitobo,Ishunju, Ruzinga, Minziro,Bugandika,Gera,Bwanjai,  Ishozi, Kanyigo, Buyango,Bugorora | Bugandika,Gera,Bwanjai,Ishozi, Kanyigo, Buyango,Bugorora |
|  | 2020 | Bukoba MC | Nyanga,Buhemba,Bilele,Bakoba,  Nshambya,Miembeni,Hamugembe,  Kashai, Kitendaguro,Ijuganyondo, Kahororo, Rwamishenye | Bilele,Bakoba,Nshambya, Miembeni,Hamugembe,Kashai, Kitendaguro,Ijuganyondo, Kahororo, Rwamishenye |
|  |  | Bukoba DC | Buhendangabo,Kanyangereko,  Maruku, Karabagaine, Kemondo, Katoma,Nyakato,Bujugo | Maruku, Karabagaine, Kemondo, Katoma,Nyakato,Bujugo |
|  |  | Missenyi DC | Buyango,Ruzinga, Gera, Bwanjai, Bugandika, Ishozi,Kanyigo | Gera, Bwanjai, Bugandika, Ishozi,Kanyigo |
|  | 2021 | Bukoba MC | Kitendaguro, Nyanga, Ijuganyondo, Rwamishenye,Bilele,Nshambya,Bakoba, Miembeni ,Kashai, Hamugembe,Kahororo | Bilele,Nshambya,Bakoba, Miembeni ,Kashai, Hamugembe,Kahororo |
|  |  | Bukoba DC | Kemondo,Bujugo, Buhendangabo, Kanyangereko,Maruku, Katoma, Karabagaine,Nyakato | Maruku,Karabagaine,Nyakato, Karabagaine,Nyakato, Katoma |
|  |  | Missenyi DC | Kitobo,Bugorora,Kashenye,Gera, Bwanjai, Kanyigo, Ishozi, Buyango,Bugandika, Ruzinga | Gera, Bwanjai, Kanyigo, Ishozi, Buyango,Bugandika, Ruzinga |
|  | 2022 | Bukoba MC | Ijuganyondo, Buhemba, Nyanga, Miembeni,Bilele,Kashai,Bakoba,  Nshambya, Kahororo,Kitendaguro | Miembeni,Bilele,Kashai,Bakoba,  Nshambya, Kahororo,Kitendaguro |
|  |  | Bukoba DC | Bujugo,Katoma, Maruku, Karabagaine | Maruku, Karabagaine |
|  |  | Missenyi DC | Bwanjai, Bugandika, Gera,Bugorora |  |
|  | 2023 | Bukoba MC | Ijuganyondo,Buhemba,Nyanga, Miembeni,Bilele,Bakoba,Nshambya,  Kashai, Kahororo,Kitendaguro. | Miembeni,Bilele,Bakoba,Nshambya,  Kashai, Kahororo,Kitendaguro. |
|  |  | Bukoba DC | Karabagaine, Maruku | Maruku |
|  |  | Kyerwa DC | Mabira |  |

*ANC = Antenatal Care, ^†^DC, district council, ^‡^CL= Confidence Level, ^§^MC, municipal council

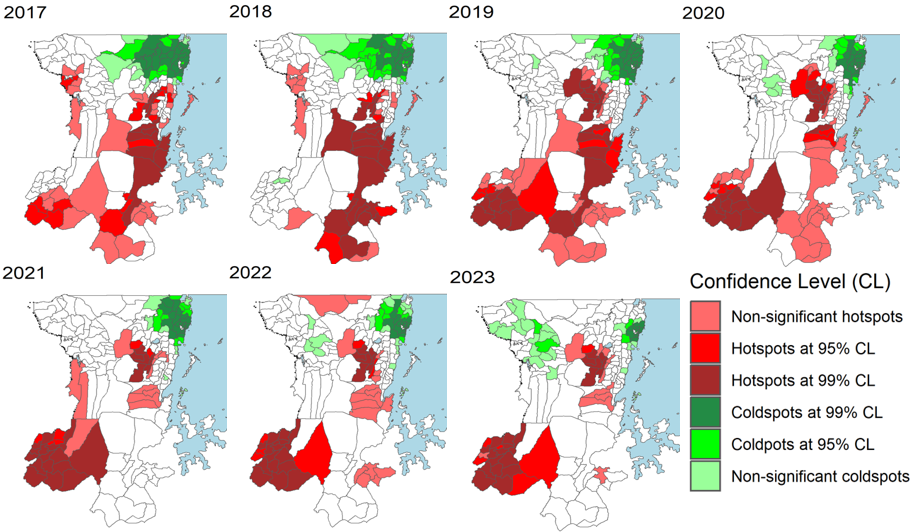
**Appendix Figure 4.** Locations of hotspots and coldspots of malaria in under-fives at ward level in Kagera region from 2017 to 2023.

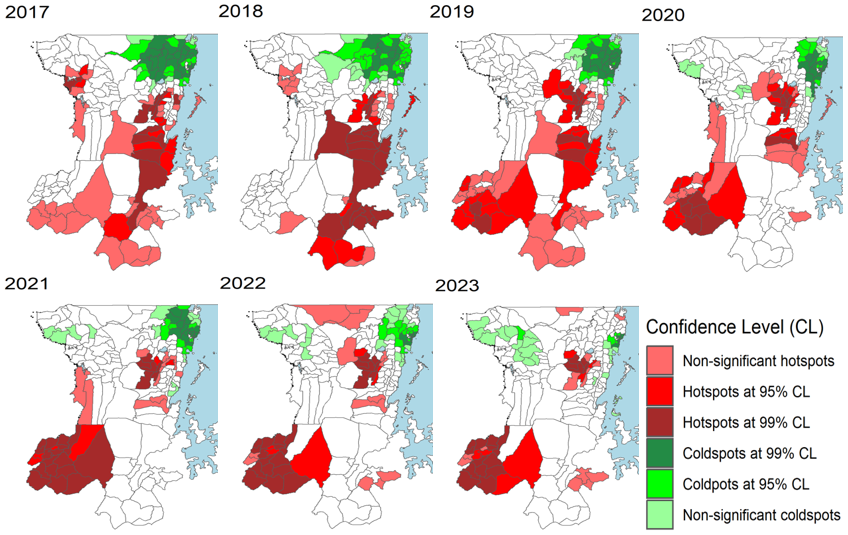

**Appendix Figure 5.** Locations of hotspots and coldspots of malaria in patients aged ≥5 years at ward level in Kagera region from 2017 to 2023.

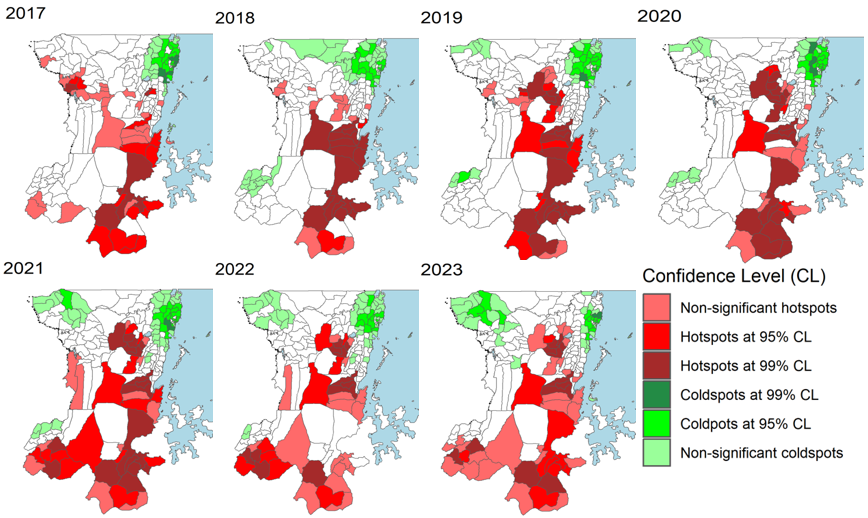

**Appendix Figure 6.** Locations of hotspots and coldspots of malaria in pregnant women at ward level in Kagera region from 2017 to 2023.

8.	Ministry of Health, Community Development, Gender, Elderly and Children, National Malaria Control Programme. Consultative Malaria Expert Meeting Report 2018. 2018.
